# The pleiotropic architecture of human impulsivity across biological scales

**DOI:** 10.1101/2023.11.28.23299133

**Authors:** Travis T. Mallard, Justin D. Tubbs, Mariela Jennings, Yingzhe Zhang, Daniel E. Gustavson, Andrew D. Grotzinger, Margaret L. Westwater, Camille M. Williams, Rebecca G. Fortgang, 23andMe Research Team, Sarah L. Elson, Pierre Fontanillas, Lea K. Davis, Armin Raznahan, Elliot M. Tucker-Drob, Karmel W. Choi, Tian Ge, Jordan W. Smoller, Abraham A. Palmer, Sandra Sanchez-Roige

## Abstract

Impulsivity is a complex psychological construct that represents a core feature of many psychiatric and neurological conditions. Here, we used multivariate methods to formally model the genetic architecture of impulsivity in humans, advancing genomic discovery and revealing pervasive pleiotropy that largely counters theories of impulsivity as a unitary construct. We identified 18 loci and 93 genes with diverse effects in GWAS and TWAS analyses, respectively, including a hotspot at 17q21.31 that harbors genes involved in neurodevelopmental and neurodegenerative disorders. Downstream analyses revealed that heterogeneous signals were localized to specific biological correlates, including expression in brain tissue during fetal development and cortical alterations in the inferior frontal gyrus. Polygenic score analyses suggested that liability for different forms of impulsivity may differentiate across development, operating via broad pathways early in life but affecting diverse outcomes by adulthood. Collectively, our study generates new insights into the pleiotropic architecture of impulsivity, which provides a more comprehensive understanding of its multi-faceted biology.

## Introduction

Impulsivity is a complex psychological construct that has a considerable role in health and behavior. Broadly characterized as a predisposition towards rapid, unplanned responses to stimuli^1^, impulsivity has been implicated in multiple psychiatric conditions^2,3^, maladaptive decision-making^4,5^, and risk-taking behaviors^6^. With well-established links to brain structure and function^4,7^, it is considered to be an endophenotype that mediates genetic effects on diverse cognitive, emotional, and behavioral domains across various clinical and non-clinical populations^3,8–12^. Given its cross-cutting relevance, a deeper etiological understanding of impulsivity could improve patient risk stratification and facilitate the development of novel therapeutic approaches with transdiagnostic applications^13–15^. However, there remains considerable debate surrounding its definition, dimensionality, and optimal measurement^1,16^, which presents a challenge to achieving these aims.

Insights from neuroscience research increasingly suggest that impulsivity can be ‘split’ into related-but-distinct facets rather than considered a unitary trait. These different aspects, such as motor and choice impulsivity, have been found to reflect only partially shared neural circuitry^7^. This recognition has spurred the development of numerous theoretical models, including reinforcement sensitivity theory^17–19^ and the dual systems model^20–22^, that attempt to explain the heterogeneous biology underlying impulsivity in humans. Although considerable progress has been made in this domain, the etiology of impulsivity remains insufficiently characterized. Disentangling shared and unique biological influences on different manifestations of impulsivity will require comprehensive multi-modal data and integrative approaches^13,23^.

In contrast to these neuroscientific findings, advances in the fields of clinical psychology, psychiatry, and genetics have revealed that many complex mental constructs can be effectively ‘lumped’ into broader categories at multiple levels of analysis. For example, multivariate models have provided compelling evidence for a hierarchical structure of mental disorders, where higher-order dimensions of psychopathology, such as thought disorder, internalizing, and externalizing, explain substantial phenotypic and genetic covariance in psychiatric diagnoses or symptoms^24–27^. This perspective has transformed our understanding of psychiatric conditions, leading to the identification of shared genetic influences, prioritizing novel transdiagnostic therapeutic targets, and accelerating genomic discovery for understudied conditions^28–33^. Considering the success of this approach in explaining the widespread pleiotropy observed among similarly complex traits^34–36^, it stands to reason that genetic studies of impulsivity may similarly benefit from lumping^37^.

With the recent completion of a large-scale genome-wide association study (GWAS) of multiple measures of impulsivity (maximum *N* = 133,517)^38^, we now have sufficient data to characterize the joint genetic architecture of multiple forms of impulsivity. In the present study, we conducted a multivariate GWAS of eight facets measured with two psychological instruments: the Barratt Impulsiveness Scale^39^ and the UPPS-P Impulsive Behavior Scale^40^ (see **Method** for further description of phenotypes). Specifically, we use the Genomic Structural Equation Modeling (Genomic SEM)^29^ framework to test the ‘splitting’ versus ‘lumping’ hypotheses with two distinct statistical models: (1) an “omnibus” model that provides an unstructured meta-analytic test of association, and (2) a common factor of impulsivity, in which associations operate through a latent genetic factor. We evaluate these models and their outputs across molecular genetic, transcriptomic, neurogenomic, and phenomic levels to gain a more comprehensive understanding of the underpinnings and correlates of impulsivity across biological scales. By doing so, we discover new insights into the complex etiology of impulsivity that advance understanding of its multifaceted nature and relationship with health and well-being.

## Results

### Genomic factor analysis reveals widespread pleiotropy among facets of impulsivity

To characterize the genomic relationships between impulsivity facets (see **Table 1** for overview), we first used linkage disequilibrium (LD) score regression to estimate the genetic covariances among study phenotypes. Generally, BIS and UPPS-P phenotypes exhibited patterns of moderate-to-large genetic overlap (mean *r*_g_ = 0.434; **Figure 1a**), except for sensation seeking which had a more modest overlap with other facets (maximum | *r*_g_ | = 0.337). Examination of the standardized genetic covariance matrix revealed that the first eigenvector explained 54% of the total genetic variance among these traits (**Figure 1b**). Consistent with this observation, we found that a common factor model provided a good fit to the data (χ^2^[17] = 325.079, CFI = 0.947, SRMR = 0.08; **Figure S1**; see **Supplementary Note** for additional details on the factor analysis). Accordingly, we proceeded with the omnibus and common factor models, as described below (**Figure 1c,d**).

**Figure 1.**
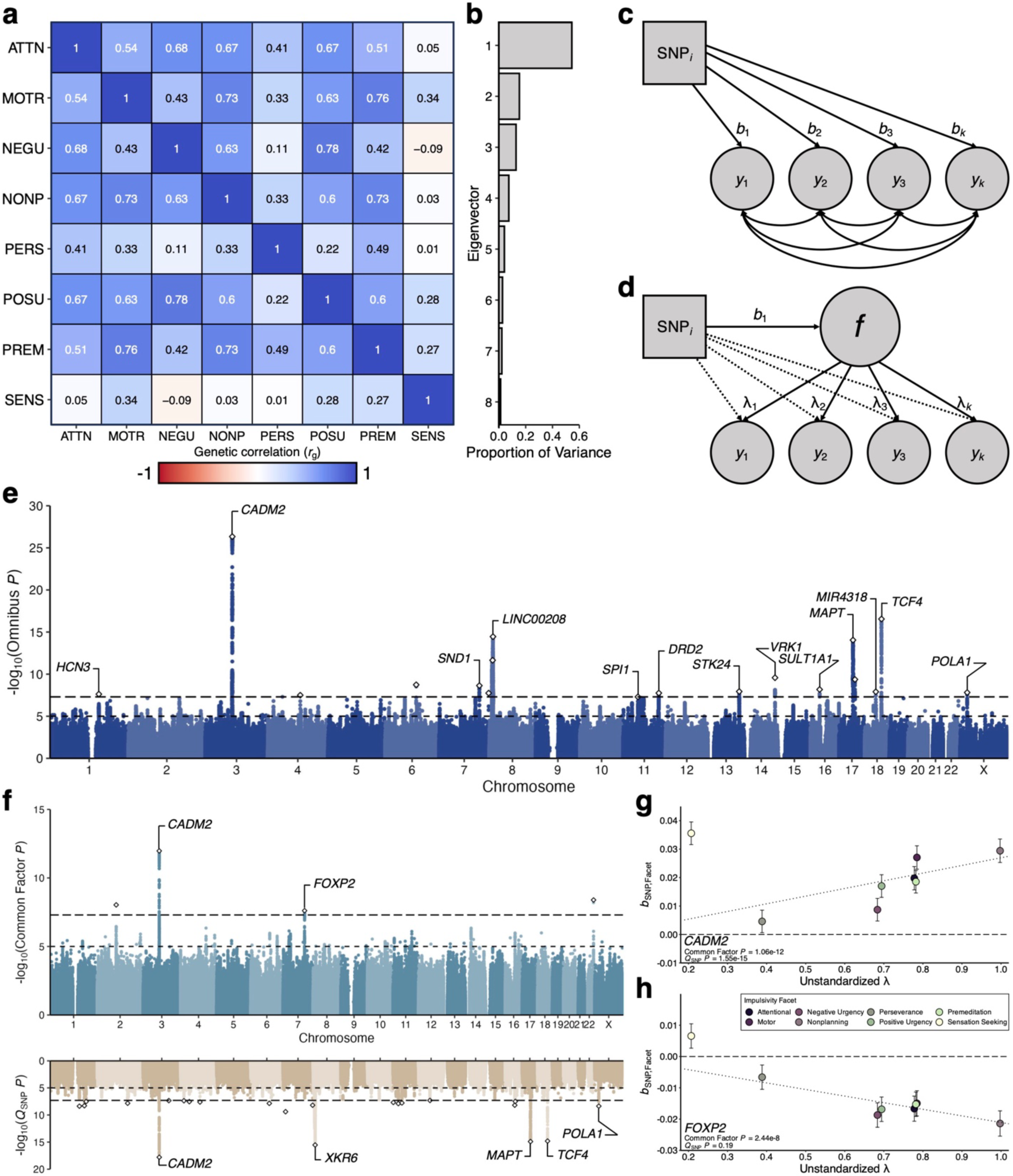
Modeling the multivariate genetic architecture of impulsivity. **a**, Matrix of genetic correlations (*r*_g_) among the eight impulsivity facets, as estimated with LD score regression. **b,** Bar chart illustrating the proportion of variance explained by each eigenvector from the genetic correlation matrix. **c,d,** Path diagrams of the two discovery models employed in the present study. The omnibus model (**c**) provides an unstructured multivariate test of association across study phenotypes (*y*_1_ - *y_k_*), while the common factor model (**d**) estimates SNP effects that operate through latent genetic factor *f*. Circles represent the inferred genetic components of model indicators and the common factor. One-headed arrows pointing from independent variables to dependent variables reflect regression relationships (regression coefficients as *b*_1_ - *b_k_*, factor loadings as λ_1_ - λ*_k_*). Dotted one-headed arrows refer to the independent pathways examined in the *Q*_SNP_ test. Two-headed arrows connecting variables represent covariance relationships. For simplicity, (residual) variances are not depicted. **e,f,** Manhattan (**e**) and Miami **(f**) plots of GWAS results for the omnibus and common factor models, respectively, where the x-axis refers to chromosomal position, the y-axis refers to statistical significance on a −log_10_ scale, the horizontal dashed line denotes genome-wide significance (*P* = 5e-8), and the horizontal dotted line marks suggestive significance (*P* = 1e-5). Lead SNPs of independent genomic risk loci are represented by white diamonds. **g,h,** Scatter plots depicting the indicator GWAS estimates and corresponding standard errors for the lead *CADM2* (**g**) and *FOXP2* (**h**) variants as a function of unstandardized factor loadings.

**Table 1.** Summary of eight impulsivity GWAS included in the present study.

| Impulsivity Facet | *N* | $h^2$ | $\lambda_{GC}$ | Mean $\chi^2$ | Intercept | Ratio | # of loci |
| --- | --- | --- | --- | --- | --- | --- | --- |
| Attentional (ATTN) | 124,739 | 0.070 | 1.147 | 1.201 | 1.026 | 0.127 | 2 |
| Motor (MOTR) | 124,104 | 0.067 | 1.147 | 1.178 | 1.011 | 0.064 | 3 |
| Negative Urgency (NEGU) | 132,559 | 0.079 | 1.200 | 1.236 | 1.017 | 0.071 | 5 |
| Nonplanning (NONP) | 123,509 | 0.093 | 1.200 | 1.252 | 1.022 | 0.087 | 6 |
| (Lack of) Perseverance (PERS) | 133,517 | 0.063 | 1.147 | 1.186 | 1.018 | 0.094 | 2 |
| Positive Urgency (POSU) | 132,132 | 0.061 | 1.147 | 1.182 | 1.017 | 0.093 | 2 |
| (Lack of) Premeditation (PREM) | 132,667 | 0.065 | 1.147 | 1.175 | 1.003 | 0.017 | 2 |
| Sensation Seeking (SENS) | 132,395 | 0.097 | 1.200 | 1.265 | 0.998 | < 0 | 7 |

Note: All GWAS summary statistics originate from Sanchez-Roige and colleagues' univariate study of impulsivity facets<sup>38</sup>. The statistics in this table were calculated with linkage disequilibrium score regression except for the number of loci, which were defined using the same pipeline as the multivariate results (**Methods**). *N* = GWAS sample size; $h^2$ = observed SNP heritability; $\lambda_{GC}$ = genomic inflation factor; mean $\chi^2$ = average $\chi^2$ statistic; intercept = estimated intercept from linkage disequilibrium score regression; ratio = the attenuation ratio, which is defined as (intercept - 1)/(mean $\chi^2$ - 1); # of loci = number of independent genome-wide significant loci.

### Multivariate GWAS and TWAS identifies novel risk genes with significant heterogeneity

Using Genomic SEM, we conducted a series of multivariate GWAS (*n*_SNPs_ = 7,053,212) and transcriptome-wide association study (TWAS; *n*_gene-tissue pairs_ = 51,698) analyses to investigate the joint genomic architecture of impulsivity in a sample of 134,935 individuals. Specifically, we employed two models for gene discovery: (1) an ‘omnibus’ model (i.e., an unstructured meta-analytic test of association) (**Figure 1c**), and (2) the common factor of impulsivity described above, where SNP- and gene-level associations operate through a latent genetic factor (**Figure 1d**). In other words, for each SNP or gene, the omnibus model tested whether the overall pattern of effects was non-zero, while the common factor model tested whether the SNP or gene influenced the indicators via a common pathway (the latent factor). *Q*_SNP_ and *Q*_Gene_ tests were also used to evaluate the degree to which effects were (in)consistent with the common pathway, providing an index of heterogeneity at each level of analysis (see **Methods** for further description and comparison of all models).

We observed substantial inflation of the GWAS test statistics for both the omnibus (λ_GC_ = 1.592, mean χ^2^ = 1.535) and common factor (λ_GC_ = 1.22, mean χ^2^ = 1.237) models (**Figure 1e,f**). The LD score regression intercepts and attenuation ratios indicated that test-statistic inflation was primarily due to robust polygenic architecture as opposed to population stratification or other confounding (omnibus: intercept = 1.021 [s.e. = 0.009], attenuation ratio = 0.039 [s.e. = 0.017]; common factor: intercept = 1.008 [s.e. = 0.008], attenuation ratio = 0.033 [s.e. = 0.034]). However, the substantially larger mean χ^2^ of the omnibus results suggests that there is an appreciable signal that is not captured by the common factor. This inference is further supported by the *Q*_SNP_ statistics (λ_GC_ = 1.607, mean χ^2^ = 1.568), which also show a stronger signal than the common factor GWAS (**Figure S2**).

With respect to the omnibus GWAS results, we identified 18 independent (*r*^2^ < 0.1) genome-wide significant loci after applying a standard clumping algorithm (**Figure 1e, Table S1**), 11 of which were novel associations that were not observed in any of the facet-level results. Some of these novel risk loci include highly pleiotropic genes, such as: a locus indexed by intronic variant rs4411372 (*P*_omnibus_ = 1.16e-8) near the *STK24* gene on chromosome 13, which has been broadly linked to cognitive and psychiatric outcomes^41–43^ a locus tagged by intergenic variant rs6507215 on chromosome 14 near *VRK1*, a gene that has been previously associated with alcohol consumption and insomnia^43,44^; and another indexed by rs6507215 (*P*_omnibus_ = 1.25e-8), an intergenic variant near *MIR4318*, which has been linked to a variety of internalizing phenotypes^45,46^. All remaining loci are reported in **Table S1**.

We found markedly fewer associated loci in the common factor GWAS. After applying a standard clumping algorithm, only four independent genomic loci were associated with a general liability toward impulsivity (**Figure 1f, Table S2**). Two signals were particularly robust: (1) rs1865250 (*b* = 0.028, s.e. = 0.004, *P* = 1.06e-12), an intronic variant in *CADM2* that was associated with several of the constituent impulsivity phenotypes and other related behaviors^38^; and (2) rs1563408 (*b* = −0.021, s.e. = 0.004, *P* = 2.44e-8), an intronic variant in *FOXP2*, a gene that has been previously linked to neurodevelopmental and externalizing conditions^31,47–49^, but not individual differences in impulsivity *per se*. Although subsequent *Q*_SNP_ tests highlighted a significant degree of heterogeneity in the *CADM2* locus (**Figure 1f,g**), the effects of the *FOXP2* variant on impulsivity facets were consistent with the common pathway model (**Figure 1f,h**). The other two genome-wide significant loci on chromosomes 2 and 22 consisted of only two SNPs, which is not indicative of a robust association (**Table S2**). Overall, these results provided limited genome-wide evidence for a common factor of impulsivity, which was further corroborated by the greater number of *Q*_SNP_ loci relative to GWAS loci (**Table S3**), as well as the finding that most *Q*_SNP_ signals were driven by multiple facets (**Figure S3**).

When examining effects at the level of gene expression via multivariate TWAS, we found a similar pattern of results. In the omnibus TWAS, we identified 325 gene-tissue pairs (88 unique genes) that were significantly associated with impulsivity (**Figure 2a, Table S4**). In contrast, no genes were significantly associated with a general liability to impulsivity in the common factor TWAS (**Figure 2b, Table S4**). Consistent with the stark contrast between our discovery models, *Q*_Gene_ analyses revealed that 96% (311/325 gene-tissue pairs) of the associations identified in the omnibus TWAS had significantly heterogeneous effects across the impulsivity facets (**Table S4**).

**Figure 2.**
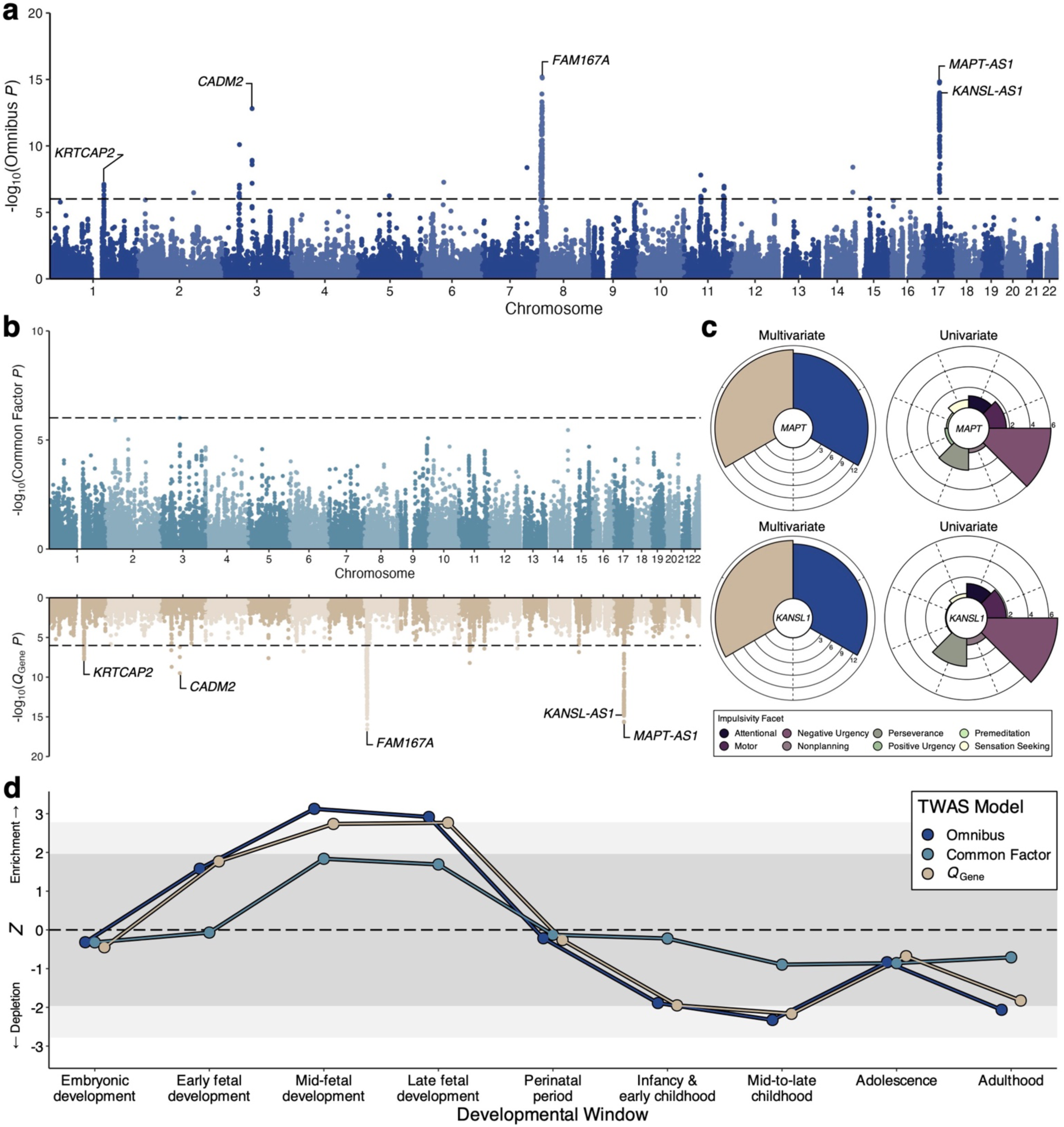
Multivariate modeling of transcriptomic architecture and its links to neurodevelopment. **a,b,** Manhattan (**a**) and Miami (**b**) plots of TWAS results for the omnibus and common factor models, respectively, where the x-axis refers to chromosomal position, the y-axis refers to statistical significance on a −log_10_ scale, the horizontal dashed line denotes genome-wide significance (*P* = 9.67e-7), and each point corresponds to one of the tested 51,698 gene-tissue pairs (spanning 13 GTEx brain tissues). **c**, Polar plots of ACAT TWAS *P* values for *MAPT* and *KANSL1* on a −log_10_ scale across all multivariate and univariate tests. **d**, Line plot of gene property analysis for TWAS results, illustrating patterns of enrichment and depletion of transcriptomic signals in developmentally specific genes across the lifespan. Developmental windows measured in PsychENCODE are plotted on the x-axis, while the *Z* statistic of enrichment or depletion is plotted on the y-axis. The dark gray band denotes the non-significant range of *Z* statistics, the light gray bands reflect nominal significance, and white reflects significance at a Bonferroni-corrected threshold.

Given the strong concordance of effects across brain tissues (all cross-tissue *r*s > 0.856), we used an aggregated Cauchy association test (ACAT) to integrate associations across all available tissues and further improve statistical power (**Methods**). The results of these multi-tissue TWAS analyses identified a total of 93, 1, and 92 unique genes associated with impulsivity in the omnibus, common factor, and *Q*_Gene_ models, respectively (**Table S5**). In the omnibus results, some of the strongest associations were concentrated within chromosome 17 at cytogenetic region 17q21.31, with *MAPT* and its non-coding RNA transcripts (*MAPT-AS1* and *MAPT-IT1*), *KANSL1* and its antisense transcript (*KANSL1-AS1*), and *CRHR1* all showing robust links to impulsivity (all ACAT *P*_omnibus_ < 7.79e-14; **Figure 2c**). Notably, these genes are located in a genetically complex segment of 17q21.31 harboring a common inversion polymorphism^50,51^ that has been linked to neurodevelopmental and neurodegenerative disorders^52–56^, as well as individual differences in brain structure^57^. For the common factor of impulsivity, the only transcriptome-wide significant association mapped to *LINC01819* (also known as *KPRT4*; ACAT *P* = 1.27e-6), a long non-coding RNA that has been associated with some psychological phenotypes (e.g., insomnia^44^) but has no known relevance to neurobiology.

### Molecular bases of impulsivity facets implicate differing aspects of neurobiology

To characterize the biological pathways involved in the etiology of impulsivity, we used MAGMA to conduct gene-based association analyses for each of the three multivariate models (**Methods**). These gene-based test statistics were then used in gene property and gene set enrichment analyses to identify specific tissues, biological processes, molecular functions, and cellular components implicated in the molecular genetic architecture of impulsivity.

Consistent with the results of our multivariate GWAS, MAGMA-based association analyses implicated 64, 1, and 68 genes in the omnibus, common factor, and *Q*_SNP_ models, respectively (**Table S6**). As expected, all three models showed significant enrichment in brain tissues, with the omnibus model evincing the broadest and strongest pattern (**Figure S4, Table S7**). We also found that the omnibus GWAS results were enriched for genes preferentially expressed during fetal neurodevelopment (*P* = 0.001; **Figure S5, Table S8**). Follow-up facet-level analyses revealed similar results. For example, all impulsivity facets showed a broad pattern of enrichment across brain tissues, albeit to a weaker degree than the multivariate models (**Figure S4**). Similarly, developmental results indicated that genes preferentially expressed during early-to-mid fetal neurodevelopment tended to be most strongly enriched among facets. The exception to this pattern involved non-planning and urgency traits, which showed the greatest relative enrichment in genes preferentially expressed during late infancy (**Figure S5**).

With respect to gene set enrichment, no gene sets were significantly enriched in the omnibus or common factor results after correcting for multiple comparisons, though we did observe an enrichment of *Q*_SNP_ signal in genes related to synapse organization (*P* = 3.39e-6). Further inspection suggested there was a general pattern of enriched signal in gene sets related to synaptic biology across models, as many of the top-ranked gene sets were related to the synapse (**Table S9**). Subsequent permutation testing indicated that synaptic location and process gene sets were indeed more strongly enriched in all GWAS results compared to other collective sets of the same size (all *P*s < 1e-4, **Figure S6**; **Methods**).

To characterize the biology implicated in the transcriptomic architecture of impulsivity, we translated the MAGMA statistical framework to facilitate gene-based association, gene property, and gene set enrichment analyses of multivariate TWAS results (**Methods**). Briefly, paralleling the results described above, we found that signal from the omnibus TWAS was enriched in genes specifically expressed during early to mid-fetal development at multiple stages (*Ps* < 0.004; **Figure 2d; Table S10**). In contrast, neither the common factor TWAS nor the *Q*_Gene_ results showed any pattern of enrichment across neurodevelopment. Facet-level results revealed that most impulsivity facets exhibited a similar pattern of enrichment as the omnibus model, except for premeditation, which showed a suggestive *depletion* of signal in genes specifically expressed during early prenatal development (**Figure S7**).

For TWAS-based gene set enrichment analyses, results implicated nitric oxide mediated signal transduction genes in the omnibus transcriptomic architecture of impulsivity (*P* = 2.92e-8; **Table S11**) – noteworthy given the key regulatory role of nitric oxide in central and peripheral neuronal signaling^58,59^. Follow-up tests revealed that this gene set was not significantly enriched in any of the lesser-powered facet-level results; however, there were nominally significant signals for attentional impulsivity and sensation seeking (*P*s = 0.003 and 0.006, respectively) that may be contributing to the omnibus signal. Finally, to parallel the MAGMA-based analyses, we performed permutation testing to evaluate whether gene sets related to synaptic locations and processes were enriched in the TWAS results, finding no evidence of categorical enrichment for these gene sets (all *P*s > 0.151; **Methods**).

### Impulsivity facets are differentially related to endophenotypes and other complex traits

Within the Genomic SEM framework, we next conducted a series of genetic correlation and *Q*_Trait_ analyses to explore how impulsivity relates to other complex traits and further evaluate the psychometric validity of the common impulsivity factor. Specifically, we used the common factor and *Q*_Trait_ models to estimate genetic relationships between impulsivity and three categories of traits: imaging-derived measures of brain structure, accelerometer-based metrics of physical activity, and psychological and psychiatric outcomes (**Methods**).

To gain greater insight into the etiological relationships between impulsivity and neuroanatomy, we estimated the genetic correlations between the common impulsivity factor and 34 bilateral regional measures of cortical thickness and surface area. The common factor was mostly negatively correlated with these measures of cortical morphology at a genetic level (**Figure 3a, Table S12**), which suggests that variants associated with a greater liability for impulsivity also tend to be associated with reduced cortical thickness (*r*_g_ = −0.21 to −0.019 [mean = −0.096]) and surface area (*r*_g_ = −0.115 to 0.063 [mean = −0.032]). Notably, some of the stronger genetic relationships between impulsivity and reduced cortical thickness were consistent with a shared pathway in frontotemporal regions (**Figure 3a,b,c**), including the lateral orbitofrontal cortex (*r*_g_ = −0.210, s.e. = 0.065, *P* = 0.001, *Q*_Trait_ *P* = 0.117) and the medial orbitofrontal cortex (*r*_g_ = −0.183, s.e. = 0.069, *P* = 0.008, *Q*_Trait_ *P* = 0.084).

**Figure 3.**
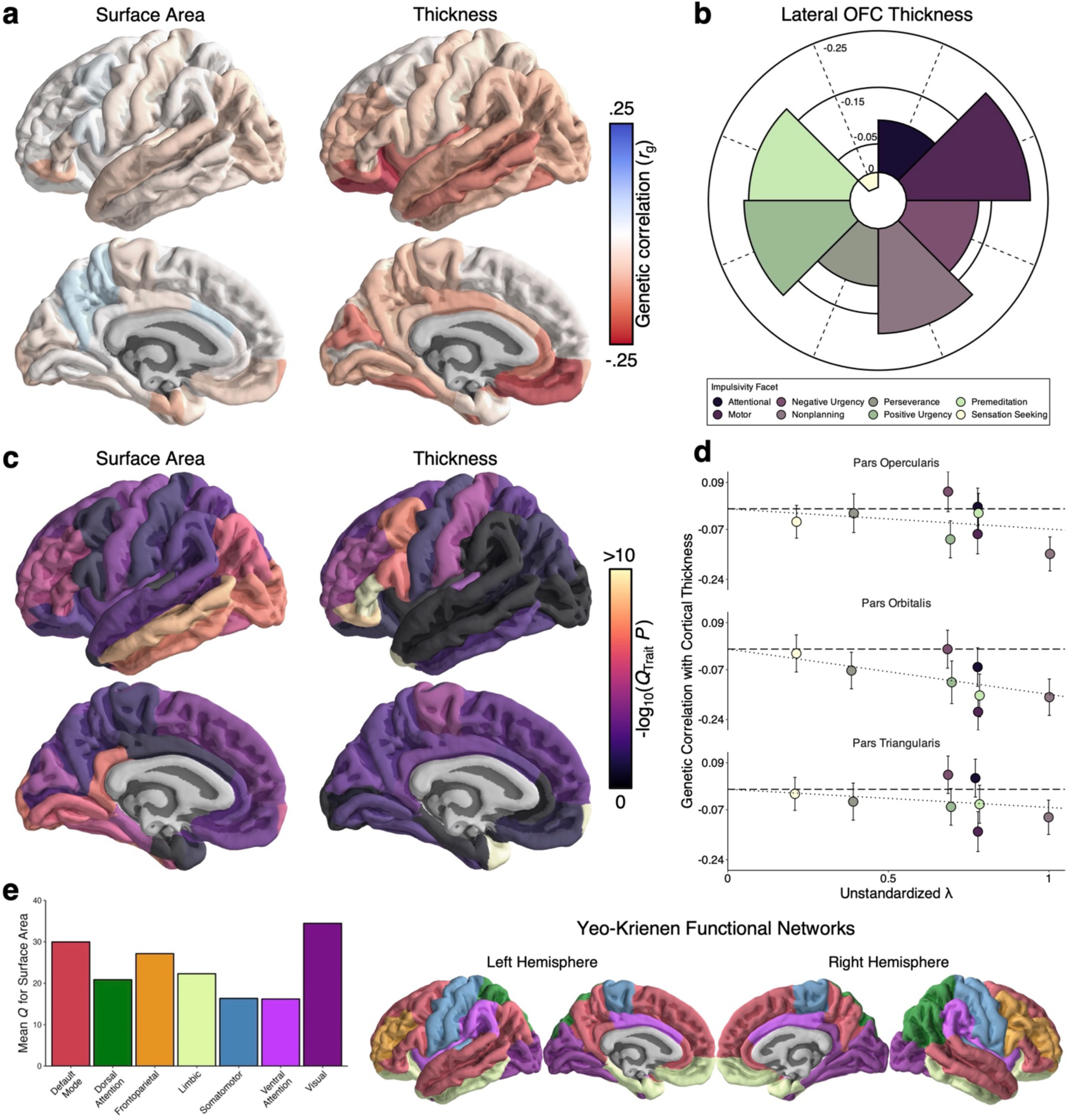
Topography of genetic relationships between cortical morphology and impulsivity. **a**, Map of genetic correlations between the common impulsivity factor and cortical surface area and thickness. **b**, Polar plot of genetic correlations between specific facets of impulsivity and thickness of the lateral orbitofrontal cortex, where facet-level effects are consistent with a shared pathway. **c**, Map of *Q*_Trait_ -log_10_(*P*) values for cortical surface area and thickness, indexing the degree to which effects are incompatible with a common factor model. **d**, Scatter plots depicting the facet-level genetic relationships for all three subdivisions of the inferior frontal gyrus as a function of unstandardized factor loadings. **e**, Bar chart illustrating the difference in *Q*_Trait_ for cortical surface area across the Yeo-Krienen functional networks, which are displayed on the right for reference.

Generally, though, *Q*_Trait_ analyses revealed widespread heterogeneity throughout the cortex for both thickness and surface area associations (**Figure 3c, Table S12**). With respect to cortical thickness, we observed strong evidence of trait-specific genetic relationships between impulsivity facets and cortical thickness, including in all three subregions of the inferior frontal gyrus: the pars orbitalis (*Q*_Trait_ *P* = 1.3e-9), pars triangularis (*Q*_Trait_ *P* = 8.09e-11), and pars opercularis (*Q*_Trait_ *P* = 2.43e-7). Notably, each of these regions has been functionally implicated in partially distinct aspects of self-regulatory control, as well as switching between goal-directed and habit-based responses. Examination of the facet-level genetic correlations revealed heterogeneity within the inferior frontal gyrus (**Figure 3d**), with *Q*_Trait_ signals driven by the divergent effects of attentional impulsivity and negative urgency, which, at times, had weak positive associations with cortical thickness. At the network level, topographical annotation indicated that cortical regions in the visual and default mode networks tended to harbor the most trait-specific surface area associations (*P*_spin_ = 0.005; **Figure 3e; Supplementary Information**).

Given that motor alterations are a prominent feature of several neurological and psychiatric disorders, we next characterized the genomic relationships between impulsivity and accelerometer-based metrics of physical activity. We found that the common impulsivity factor was modestly genetically correlated with physical activity across the day, with positive genetic correlations during the early morning hours (00:00–06:59) and negative genetic correlations throughout the remainder of the day (07:00–23:59) (**Table S13; Figure S8**). Only 3 hourly associations were significant following correction for multiple testing, all of which showed significant evidence of heterogeneity in subsequent *Q*_Trait_ analyses. Inspection of the facet-level genetic correlations revealed that the links between impulsivity and physical activity were generally similar during morning hours, but these diverged in the afternoon and evening (**Figure 4a, Table S13**). Perseverance and sensation seeking displayed some of the clearest facet-specific patterns, with the former linked to a more pronounced dip in activity during the afternoon (lowest *r*_g_ = −0.231) and the latter linked to increased activity during the evening (highest *r*_g_ = 0.178).

**Figure 4.**
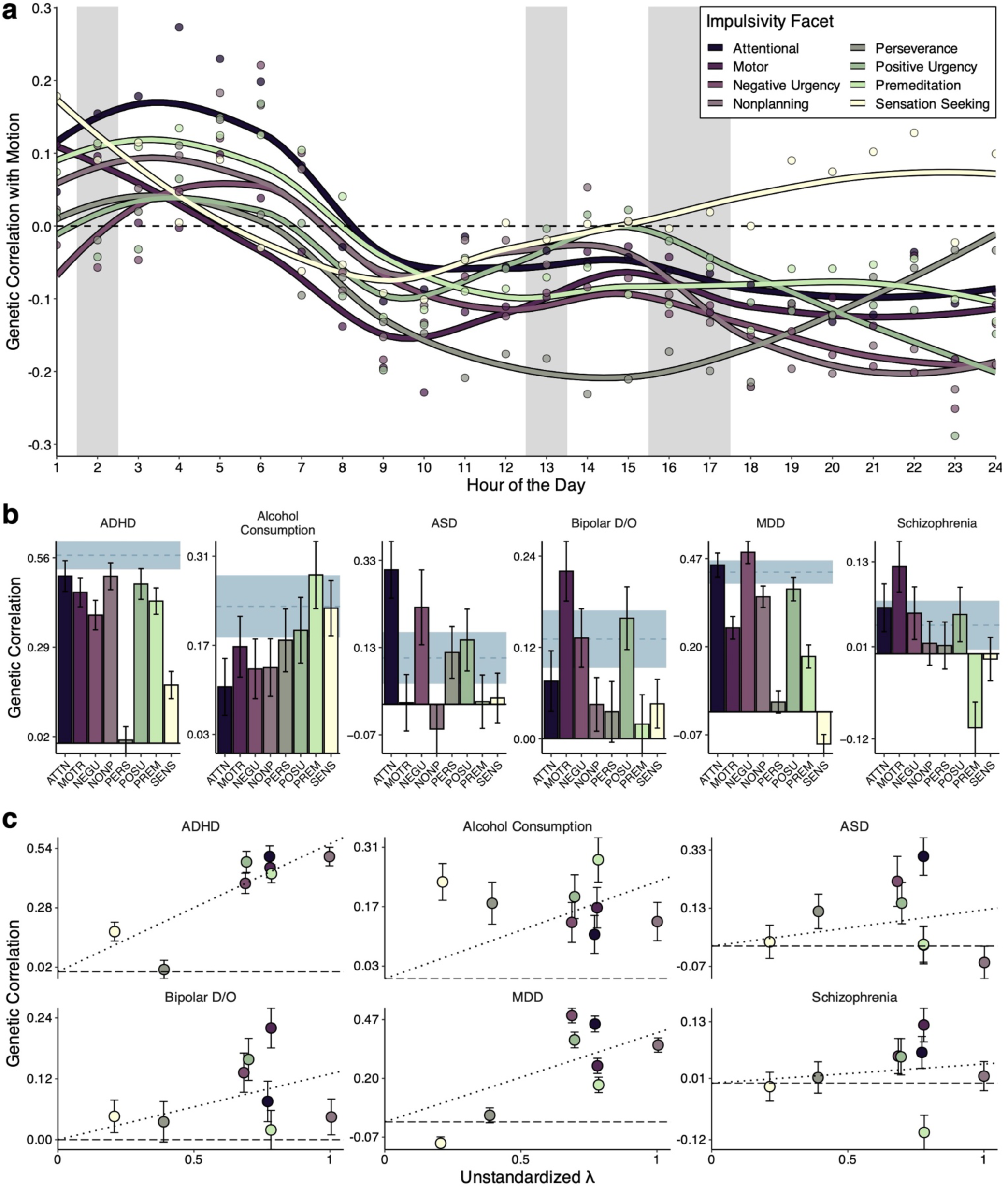
Facet-specific and heterogeneous genetic correlations between impulsivity and behavioral phenotypes. **a**, Line plots of genetic correlations between impulsivity facets and accelerometer-based measures of physical activity, as estimated with LOESS. Vertical gray bands highlight time periods when *<u>Q</u>*_Trait_ *P* was non-significant (i.e., periods when the observed pattern of effects was consistent with a common pathway). **b**, Bar charts depicting the genetic correlations between impulsivity facets and select psychiatric outcomes. Horizontal blue lines index the genetic correlation between an outcome and the impulsivity factor (and the corresponding standard error). **c**, Scatter plots illustrating how genetic correlations with psychiatric outcomes are inconsistent with a common factor model in multiple ways, where facet-level relationships are plotted as a function of unstandardized factor loadings. Error bars depict standard errors.

Finally, we examined genomic relationships between impulsivity and 23 psychiatric phenotypes. As might be expected, genetic correlations between the common impulsivity factor and these outcomes were generally positive (range = 0.039 - 0.621) and statistically significant (**Figure 4b, Figure S9)**. We observed some of the largest genetic correlations with attention-deficit/hyperactivity disorder (*r*_g_ = 0.567, s.e. = 0.043), alcohol use disorder (*r*_g_ = 0.535, s.e. = 0.072), and cannabis use disorder (*r*_g_ = 0.479, s.e. = 0.058). However, follow-up *Q*_Trait_ analyses revealed significant facet-level specificity for every tested trait (all *Q*_Trait_ *P* < 8.48e-6; **Figure 4c, Table S14; Figure S10**). For example, attention-deficit/hyperactivity disorder and its various clinical presentations (e.g., childhood, persistent, late-onset) exhibited null genetic correlations with perseverance in a manner that deviated from the common pathway model. Other notable deviations include facet-specific genetic correlations between schizophrenia and premeditation (*r*_g_ = −0.104, s.e. = 0.036), bipolar disorder and motor impulsivity (*r*_g_ = 0.22, s.e. = 0.039), and alcohol consumption and sensation seeking (*r*_g_ = 0.228, s.e. = 0.043) (**Figure 4c**). Collectively, results revealed that genetic relationships with psychopathology were inconsistent with a model of general liability to impulsivity and instead better captured with facet-specific pathways.

### Facet-level polygenic scores are generally more informative than one for a common factor of impulsivity

We next tested whether a polygenic score for common impulsivity might be more strongly or broadly associated with impulsivity and other health-related outcomes. To accomplish this, we calculated polygenic scores for the common factor of impulsivity and the constituent facets in two independent samples that span most of the human lifecourse: the Adolescent Brain and Cognitive Development Study (ABCD)^60,61^ and the Vanderbilt University Medical Center biobank (BioVU)^62,63^. ABCD was used to examine associations between polygenic scores and psychological measures, while BioVU was used to examine associations with medical outcomes.

In ABCD, we found that the common factor polygenic score was robustly associated with psychological measures closely related to impulsivity, with similar Δ*R*^2^ values as the facet-level polygenic scores across eight outcomes: the five UPPS-P subscales (negative urgency, [lack of] premeditation, [lack of] perseverance, positive urgency, and sensation seeking) and three subscales from the Child Behavior Checklist (CBCL)^64,65^ (attention, externalizing, and internalizing problems). Here, through a set of novel *Q*-like analyses, we found that the effects of facet-level polygenic scores largely aligned with a common pathway model (**Figure 5a, Methods**) – a finding that was further supported by the relatively strong performance of the impulsivity factor polygenic score in ABCD (**Table S15**). There were, however, six instances where the effects of sensation seeking or perseverance polygenic scores significantly deviated from expectation (**Table S15**, **Figure S11**), suggesting that some relationships in the sample are better captured by facet-specific versus shared pathways.

**Figure 5.**
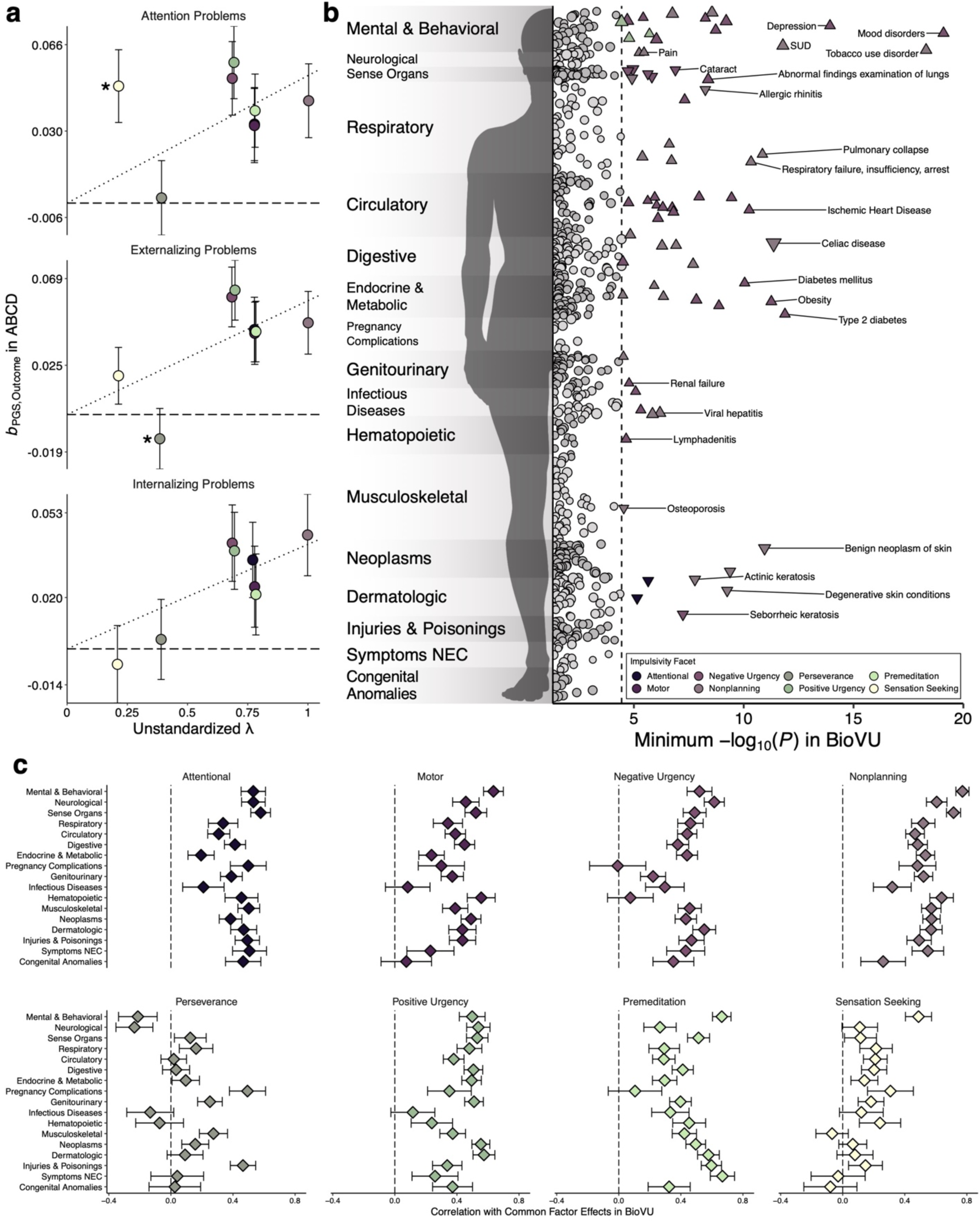
Polygenic scores and the validity of a common pathway in independent datasets. **a**, Scatter plot depicting the effect of each facet-level polygenic score for the CBCL outcomes in the ABCD Study as a function of the unstandardized factor loadings. Error bars depict the standard error. Asterisks denote significant deviations from the common pathways model. **b**, Manhattan plot of the strongest associations between facet-level polygenic scores and medical outcomes defined by “phecodes” in BioVU. The y-axis refers to the category of phecode, the x-axis refers to the statistical significance of the association on - log10 scale, and the dashed line denotes the Bonferroni-adjusted significance threshold (*P* = 0.05/1,380 = 3.62e-5). Each point in the plot corresponds to the minimum *P* value for an association between the eight facet-level polygenic scores and a phecode. Circles denote nonsignificant associations, upward-facing triangles are significant positive associations, and downward-facing triangles are significant negative associations. Size reflects the effect size, with larger points reflecting a larger absolute effect. For plotting purposes, the x-axis has been truncated to begin at *P* < .05 (1.301 on the -log10 scale), and conditions that did not meet this threshold were omitted. NEC = not elsewhere classified, SUD = substance use disorder. **c**, Scatter plot of the correlation between the effects of the common factor polygenic score and the effects of facet-level polygenic scores, where error bars correspond to the standard error of the estimate.

A very different pattern emerged in BioVU, where the common factor polygenic score was neither strongly nor broadly associated with health-related outcomes, with only five phenome-wide significant associations. By comparison, polygenic scores for negative urgency and nonplanning were linked to 52 and 47 medical outcomes, respectively. More precisely, when compared to all facet-level polygenic scores, significant associations with 67 unique health outcomes were obfuscated when the common factor polygenic score was used (**Figure 5b, Table S16**, **Figures S12-20**). Some of the missed relationships included novel links between urgency traits and increased odds for neurodevelopmental disorders (e.g., negative urgency–autism spectrum disorder: OR = 1.219, *P* = 2e-8; positive urgency–autism spectrum disorder: OR = 1.197, P = 8.82e-5; positive urgency– pervasive developmental disorder: OR = 1.119, *P* = 1.64e-5), negative urgency and increased odds for disorders of the circulatory system (e.g., negative urgency–myocardial infarction: OR = 1.103, *P* = 1.57e-7), and nonplanning and increased odds for viral hepatitis C (OR = 1.151, *P* = 1.42e-6). In addition to the expected associations with externalizing disorders (e.g., substance use disorders), there were significant positive links between multiple facets of impulsivity and internalizing conditions (e.g., major depressive disorder), buttressing the etiological connections between a broad spectrum of dysregulated behavioral and emotional processes.

Given the distinct PheWAS results, we sought to better understand the relationship between the common factor and facet-level polygenic score effects. In general, we found that the correlation between the two varied considerably across scores and medical categories (**Figure 5c**). Even in the mental disorders category, where concordance was generally higher, patterns ranged from modest negative correlations (*r* = −0.213 for perseverance) to strong positive correlations (*r* = 0.771 for nonplanning impulsivity).

## Discussion

Advancing etiological understanding of impulsivity is necessary to develop more effective therapeutic interventions for numerous psychiatric and neurological disorders. In the present study, we investigated the genetic architecture of impulsivity, employing complementary multivariate models to advance genomic discovery and formally evaluate pleiotropy across biological scales. Our results provide insight into the heterogeneous biology underlying impulsivity, revealing pervasive pleiotropy that is largely inconsistent with a common factor model of impulsivity in downstream analyses at molecular genetic, transcriptomic, neurogenomic, and phenomic levels. Three key insights emerge from this work: (1) there is more evidence for *splitting* over *lumping* that emerges at granular levels of analysis, especially for sensation seeking and (lack of) perseverance; (2) despite limited support for lumping, there are instances where the common factor approach may facilitate robust and interpretable discoveries for impulsivity; and (3) the etiology of impulsivity is closely linked to early neurodevelopment and related conditions.

The considerably greater evidence for fractionating impulsivity stands in stark contrast to recent findings in psychiatric and behavioral genetics, which indicate that substantial genetic risk is shared across related psychological outcomes. This is particularly striking since a common factor model showed good fit to our data, and this would typically suggest overlapping architecture that is consistent with a general dimension of liability. However, while a parsimonious lumping approach seemed viable at the level of genome-wide covariance, it performed poorly in subsequent analyses where many trait-specific patterns and signals were observed – especially for sensation seeking and (lack of) perseverance. These results align with related insights from neuroscience^7^, and they underscore the importance of evaluating conceptual models across levels of analysis, especially considering the growing trend to group related psychological constructs together^13,14^.

Notably, our results show a distinct advantage of the omnibus model (i.e., the unstructured splitting approach) in GWAS and TWAS results. This approach markedly improved statistical power, enabling the discovery of novel risk loci and biological pathways related to multiple aspects of human impulsivity. For instance, our results revealed strong novel signals within chromosome 17 that localized to cytogenetic region 17q21.31. Here, we found a particularly robust association between impulsivity and the *MAPT* gene, along with its related non-coding RNA transcripts *MAPT-AS1* (antisense) and *MAPT-IT1* (intronic). Converging lines of evidence have linked genetic variation in *MAPT* to difficulties with self-regulation in the general population^31,66^, with potential mechanisms including altered expression of either MAPT or its regulatory RNA transcripts, which can disrupt tau protein function^67^. This disruption can lead to aberrant microtubule dynamics, synaptic dysfunction, and compromised neural circuitry involved in impulse control and executive functions^68,69^. We also identified robust associations between impulsivity and *KANSL1* and its antisense transcript, *KANSL1-AS1*, which play a key role in autophagy and synaptic activity^70^. Variation in this gene has previously been linked to neurodevelopmental and neurodegenerative disorders that exhibit difficulties with self-regulation^53,56,71^, and changes to its expression or function could influence individual differences in impulsivity as well.

The improved statistical power of the omnibus model also facilitated new discoveries in downstream analyses, such as links to synaptic biology and nitric oxide signaling in the genomic and transcriptomic architecture of impulsivity, respectively. Nitric oxide plays a complex and multifaceted role in the central nervous system^59^, where it is involved in mediating and modulating neurotransmission, neurovascular coupling, and neuroinflammatory processes. Disrupted nitric oxide metabolism also engenders impulsive, hyperactive, and aggressive behavior in murine models^72–75^, and altered nitric oxide signaling is linked to attention deficits in patients with certain neurometabolic disorders (e.g., argininosuccinate lyase deficiency)^76^.

Our heterogeneity tests also advanced understanding of the pleiotropy across biological scales by identifying loci with divergent or trait-specific effects, such as *CADM2*. Here, we used *Q*_SNP_ and *Q*_Gene_ analyses to reveal that the effects of *CADM2* were highly specific to certain facets and inconsistent with a general liability to impulsivity, granting new insights not afforded by univariate GWAS or TWAS^38^. Bioinformatic analyses indicated that the most heterogeneous and facet-specific signals localized to genes that are preferentially expressed during early neurodevelopment. Furthermore, our *Q*_Trait_ analyses detected several divergent effects on brain structure. We found that the inferior frontal gyrus exhibited pronounced heterogeneity with impulsivity, with highly facet-specific signals observed in all three of its cytoarchitecturally diverse subdivisions: the pars orbitalis, pars triangularis, and pars opercularis. As alterations in cortical thickness partly reflect variability in cellular organization (e.g., disturbances in neural progenitor cell proliferation or neuronal differentiation)^77,78^, differences in cell type distributions or morphology may influence regional-specific links to impulse control and decision-making.

Beyond clarifying the unique biology associated with specific impulsivity facets, the *Q* tests also allowed us to identify effects that plausibly operate via a common pathway, such as the *FOXP2* gene. *Q*_SNP_ and *Q*_Gene_ results indicated that the effects of this gene on impulsivity were consistent with a general liability model, which is noteworthy given its prior associations with externalizing psychopathology^31,48,49^, as well as speech and language disorders^47,79,80^. With established roles in the regulation of neural plasticity, synaptic connectivity, and neurite outgrowth^81^, *FOXP2* may exert a highly pleiotropic effect on psychological outcomes during early development, with impulsivity serving as an intermediate phenotype. Furthermore, we found several nominal genetic correlations with cortical thickness that were consistent with a common pathway model, such as the medial and the lateral orbitofrontal cortex (OFC). Both of these regions have been functionally implicated in impulsive behavior, with medial OFC responses encoding the subjective value of delayed reward in order to support goal-directed behavior^82^, and lateral OFC responses, together with the insula, supporting learning from aversive outcomes to guide adaptive behavior^83–85^.

Finally, we identified several specific risk genes and pathways that are involved in critical developmental programs, and we found that both the genetic and transcriptomic architectures of impulsivity are enriched for genes that are primarily expressed during early-to-mid fetal neurodevelopment. These links were underscored in our polygenic score analyses, including the first PheWAS of impulsivity, where we identified novel associations between facet-level polygenic scores and neurodevelopmental conditions. In scanning the medical phenome, we found that genetic liability for urgency-related traits was linked to autism spectrum disorder in a manner that appeared to be trait-specific, with effects that were larger than any other association (e.g., negative urgency– autism spectrum disorder: OR = 1.219; positive urgency–autism spectrum disorder: OR = 1.197). Notably, these relationships were not observed when using a common factor polygenic score. However, although the common factor polygenic score was not informative in adults, we did find that it was robustly associated with psychological measures in children, where facet-level effects largely aligned with a common pathway model. One interpretation of this pattern is that genetic risk for impulsivity largely operates via broad, general pathways in children but canalizes into more unique channels across development, such that a general index becomes less informative by adulthood, as seen for problematic alcohol use^86–88^.

These findings should be interpreted in light of several limitations, such as the exclusion of individuals with non-European ancestry due to methodological constraints. This, coupled with the participation biases in the present cohort^89^, may affect the generalizability of our findings to other populations. Furthermore, all univariate GWAS analyses were conducted using facet-level sum scores from the BIS-11 and the UPPS-P, as opposed to item-level measures. It has previously been demonstrated that the use of item-level modeling can ameliorate certain confounds and generate deeper insights into the shared genetic architecture among complex traits^30,66,90^. Additionally, the present study was limited to self-report measures of impulsivity, which may not fully generalize to task-based measures of similar cognitive and behavioral processes. Thus, future research should seek to replicate and extend our findings in more diverse and representative samples with more granular measures of impulsive traits.

Nevertheless, our study considerably advances understanding of impulsive traits and their genetic architectures. Through multivariate modeling across biological scales, we identified novel risk genes for impulsivity facets, uncovered trait-specific effects at multiple levels of analysis, and highlighted new genetic links with neurodevelopment. While we do find several instances of convergent biology that implicate a shared pathway, we find substantially more evidence for trait-specific biology underlying the impulsivity facets. Sensation seeking and perseverance clearly differ from other facets in their etiological underpinnings, and they are perhaps better conceptualized as indices of psychological motivation or reward sensitivity rather than self-regulation, per se^37^. Indeed, this would align with phenotypic factor analytic research that suggests these facets tend to be more distinct from other impulsive traits^91,92^.

Collectively, these results cast considerable doubt on the validity of impulsivity as a unitary construct, challenging the conventional practice of lumping together diverse cognitive and behavioral tendencies that share only modest overlap. Instead, our findings advocate for a more refined and nuanced approach to studying impulsivity and its links to psychiatric and neurological disorders, one that recognizes its multifaceted nature and the distinct biology that influences its various facets. This shift in perspective paves the way for a deeper exploration of impulsive traits, promising new insights into the complexities of human psychology and health.

## Methods

This study is accompanied by **Supplementary Information**, which includes additional tables and figures.

### GWAS data acquisition and pre-processing

Univariate GWAS summary statistics were obtained from a previously published study of impulsivity^38^. In the present study, we analyzed GWAS summary statistics for two psychometrically validated measures of impulsivity: the Barratt Impulsiveness Scale (BIS-11)^39^ and the UPPS-P Impulsive Behavior Scale^40^. The former includes subscales that measure attentional, motor, and nonplanning impulsiveness, while the latter includes subscales that measure (lack of) premeditation, (lack of) perseverance, positive urgency, negative urgency, and sensation seeking. These phenotypes are succinctly described below:

- **Attentional Impulsiveness (ATTN)**: An inability to concentrate or focus attention.
- **Motor Impulsiveness (MOTR)**: A predisposition to hastily engage in behaviors in response to internal or external stimuli.
- **Negative Urgency (NEGU)**: A tendency to react rashly to negative affect without consideration of potential consequences.
- **Nonplanning Impulsiveness (NONP)**: An inclination to forgo future planning and forethought.
- **Lack of Perseverance (PERS)**: An inability to remain focused on tasks that may be boring or difficult.
- **Positive Urgency (POSU)**: A tendency to rashly react to positive affect without consideration of potential consequences.
- **Lack of Premeditation (PREM)**: A tendency not to think about or reflect on the consequences of an action before taking it.
- **Sensation Seeking (SENS)**: A predisposition to seek out new and exciting experiences, potentially without evaluating the risk.

As described by the original authors^38^, these GWAS were conducted in a cohort of up to 133,517 participants of European ancestry collected by 23andMe, Inc., a direct-to-consumer genetics company. Further description of the cohort can be found in the original study by Sanchez-Roige and colleagues^38^.All summary statistics included in the present analysis had significant SNP-based heritability and robust polygenic signal (**Table 1**). Reference alleles were aligned across univariate GWAS summary statistics, and only SNPs with a minor allele frequency ≥ 0.5% and an imputation quality score ≥ 0.6 were included in analyses.

### Multivariate GWAS of impulsivity

Using Genomic SEM v0.0.5^29,33^, we performed multivariate genome-wide association analyses for two distinct statistical models: the omnibus model (i.e., “splitting” impulsivity) and the common factor model (i.e., “lumping” impulsivity). We also perform follow-up *Q*_SNP_ tests to determine whether SNP effects on the common factor are statistically plausible. Details of the exploratory and confirmatory factor analyses are reported in the **Supplementary Information**. All analyses were conducted adhering to best practices, using standard reference panels and parameter settings, and diagonally weighted least squares (DWLS) estimation. The estimated sample size of the latent genetic factor was calculated in the manner described by Mallard and colleagues^32^.

We refer interested readers to the original methodology papers by Grotzinger and colleagues^29,33^ for exhaustive description of the method, but we provide an overview of the genomic structural equation models reported herein below. As illustrated in **Figure 1c,d**, the omnibus model tests the degree to which a SNP has non-zero effects on impulsivity facets, the common factor model tests the degree to which a SNP influences impulsivity facets via a common pathway, and the *Q*_SNP_ model tests the degree to which said common pathway is statistically plausible.

#### Omnibus model

To test whether the overall pattern of effects for a given SNP was non-zero across study phenotypes, we performed genome-wide unstructured tests of association, where all relationships among impulsivity facets and their relationships with tested SNPs were freely estimated. This was accomplished by first fitting the following system of regressions for each SNP:

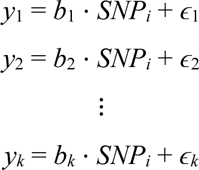

Where *y* refers to the *k* facets in the structural equation model, *SNP_i_* is the tested SNP, *b* represents the estimated regression coefficients, and *ɛ* denotes the error terms. We then fit a null model with all regression coefficients constrained to equal zero for the tested SNP, such that

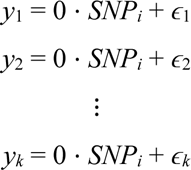

The difference in fit between these two models was then calculated with a *χ*^2^ difference test, where Δ*χ*^2^ = *χ*^2^_free_ *- χ*^2^_constrained_ and Δ*df* = *df*_free_ *- df*_constrained_. The omnibus *P* value for *SNP_i_* can then be calculated as *P* = 1 - *CDF*_*χ*^2^_ (Δ*χ*^2^, Δ*df*), where *CDF*_*χ*^2^_ is the cumulative distribution function of the chi-square distribution with Δ*df* degrees of freedom.

#### Common factor model

To estimate the effect of a given SNP on a general dimension of impulsivity, we used the common factor approach described by Grotzinger and colleagues^29^ to model the genetic covariances among study phenotypes. In this framework, a measurement model describes the genetic relationships between model indicators and a latent factor as

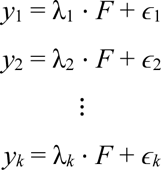

Where *y* refers to the *k* observed indicators in the measurement model, *F* is the latent genetic factor, λ represents the factor loadings, and *ɛ* denotes the residual variances or errors associated with each indicator. The latent genetic factor can then be regressed onto a given SNP as

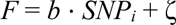

Here, *b* refers to the regression coefficient representing the effect of *SNP_i_* on *F*, and ζ is the residual. Note that the standard error and *P* value for this estimated parameter were computed within the model, as well.

#### Q_SNP_ model

To statistically test whether the estimated SNP effect plausibly operated via the latent factor (i.e., a common pathway), we conducted follow-up *Q*_SNP_ tests for every tested SNP. This involved comparing the common factor model to one where SNPs directly influence indicators through freely estimated independent pathways, as opposed to the common pathway. The difference in model fit can then be quantified with a *χ*^2^ difference test, where Δ*χ*^2^ = *χ*^2^_independent_ *- χ*^2^_common_ and Δ*df* = *df*_independent_ *- df*_common_. The *Q*_SNP_ *P* value for *SNP_i_* was subsequently calculated as *P* = 1 - *CDF*_*χ*^2^_ (Δ*χ*^2^, Δ*df*), as described above.

Note that the expectation is not necessarily that SNP-level effects are homogenous across model indicators. Instead, under the common pathway model, we would expect that the effect of a given SNP on model indicators scales proportionally with the unstandardized factor loadings, where

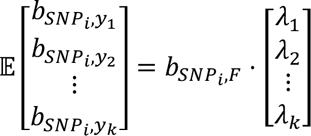

Here, *b_SNP_i_,y_k__* represents the effect of SNP *i* on indicator *k*, *b_SNP_i_,F_* is the effect of the SNP *i* on latent factor *F*, and λ*_k_* represents the unstandardized loadings on the latent factor. As described by de la Fuente and colleagues^93^, misfit will occur when the vector of expected SNP effects on indicators is discordant with the observed SNP effect on indicators. The resulting *Q* statistic will be high, and the model in which the SNP influences indicators exclusively via the common pathway will be rejected. Interpretation can be facilitated by plotting the univariate GWAS betas by the unstandardized factor loadings, as illustrated in **Figure 1g,h**.

### Biological annotation of GWAS results

To characterize genome-wide significant loci, we used the FUMA v1.5.4 SNP2GENE pipeline with default parameters to apply a standard clumping algorithm to our GWAS results. The European ancestry samples from the 1000 Genomes Project Phase 3v5 (*n* = 503) were used as a LD reference panel. To identify putative risk genes, we conducted positional mapping using field standard ANNOVAR annotations (version 2017.07.17). We also used FUMA to investigate whether loci associated with impulsivity have been previously linked to other complex traits in the GWAS Catalog (version e0_r2022-11-29).

#### Gene-based analyses

We used MAGMA^94^ v1.08 to conduct gene-based association, gene property, and gene set enrichment analyses. Standard procedures were followed for gene-based association analyses based on GWAS summary statistics and default MAGMA parameters were employed throughout the pipeline.

MAGMA was also used to conduct competitive gene property and gene-set enrichment analyses based on the gene-level *P* values produced in the association analyses. These analyses evaluated whether genes in a specific annotation had a stronger association with the GWAS outcome compared to other genes.

In the gene property analyses, we utilized the GTEx v8 dataset, comprising 54 tissues, to examine enrichment in various bodily tissues. Additionally, we used brain tissue data from 11 developmental epochs in the BrainSpan dataset to assess enrichment throughout neurodevelopment. In the gene set enrichment analyses, we tested up to 9,988 gene sets from the Molecular Signatures Database v2023.1. These sets correspond to 7,343 biological processes, 1,001 cellular components, and 1,644 molecular functions. For determining statistical significance, we used Bonferroni-corrected thresholds of *P* ≤ 9.26e-4 for the GTEx tests, *P* ≤ 4.54e-3 for the BrainSpan tests, and *P* ≤ 5.01e-6 for the gene set tests.

#### Permutation test for synaptic gene sets

Given the relatively strong signals observed for several gene sets related to the synapse, we sought to test whether gene sets robustly implicated in synaptic biology, especially those robustly linked to synaptic locations and processes, were significantly enriched in the genetic architecture of impulsivity. To accomplish this, we utilized the SynGO database^95^, a meticulously curated repository of synaptic gene ontologies, to identify with high confidence gene sets related to the synapse as well as those that were not. We then used a permutation-based approach to test whether the mean *Z* statistic from our synapse-related gene sets was significantly different from what we might expect by chance, which we empirically established by randomly sampling the mean *Z* statistic of 10,000 combinations of gene sets of the same size. Statistical significance was calculated based on the number of times the observed value was greater than the randomly sampled value (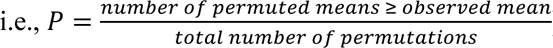).

### Multivariate TWAS of impulsivity

To investigate the effects of tissue-specific gene expression on impulsivity, we used Transcriptome-Wide Structural Equation Modeling^96^ (version 0.0.5), which is based on the FUSION TWAS method. Specifically, we used this approach to conduct multivariate transcriptome-wide association analyses for the omnibus and common factor models. We also performed follow-up tests and calculated *Q*_Gene_ statistics to evaluate whether SNP effects on the common factor were plausible.

Within this framework, GWAS summary statistics for the impulsivity facets were then paired with *cis* gene expression quantitative trait locus data for 13 brain tissues from the Genotype-Tissue Expression Project (GTEx) v8 dataset to produce univariate TWAS estimates for 51,698 gene-tissue pairs. These gene-level summary statistics were then used as input for the multivariate TWAS, which yielded results for the omnibus, common factor, and *Q*_Gene_ models.

#### Aggregated Cauchy association test

Given that effects were strongly and positively correlated across tissues (all cross-tissue *r*s > 0.856), we used an aggregated Cauchy association test (ACAT) to integrate information across all available tissues and improve statistical power prior to downstream bioinformatic analyses. Specifically, for each multivariate model, we used the ACAT R package (version 0.91) to combine TWAS *P* values across available tissues for each gene. Following the recommendations of the developers, any ACAT *P* value equal to 1 was replaced by 1-1/*d*, where *d* refers to the number of TWAS *P* values combined by ACAT. We calculated *Z* statistics using the inverse cumulative density function (or quantile function) of the standard normal distribution, such that 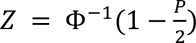.

### Biological annotation of TWAS results

To assess the enrichment of TWAS signals across various biological contexts, we employed a series of regression-based procedures, drawing inspiration from the MAGMA framework^94^. Specifically, we examined enrichment across neurodevelopmental epochs and gene ontology terms.

For neurodevelopmental enrichment, we leveraged gene expression data from the PsychENCODE Consortium, deriving gene expression specificity scores for nine neurodevelopmental epochs (see following subsection). Each epoch’s enrichment was then assessed by regressing gene-level *Z* statistics, obtained from the ACAT TWAS *P* values, on specificity scores for each developmental window. The median number of SNPs used in the TWAS weights, the median gene expression heritability, and the number of TWAS *P* values combined via ACAT were included as covariates in this analysis.

For gene ontology terms, we characterized the enrichment of TWAS signals for all terms in the Molecular Signatures Database v2023.1. This was achieved by regressing the ACAT TWAS *Z* statistics onto a binary variable indicating gene set membership while accounting for the same gene-level covariates described above. Gene sets with less than three genes available for analysis were excluded. Finally, to parallel our GWAS bioinformatic analyses, we also conducted permutation-based tests of enrichment for SynGO^95^ gene sets as described in the previous section.

Statistical significance was tested using two-sided *P* values for the continuous neurodevelopmental specificity scores, while one-sided *P* values were used for the binary gene sets, consistent with the MAGMA framework. Bonferroni-corrected thresholds for significance were set at *P* ≤ 5.55e-3 for developmental analyses and *P* ≤ 7.72e-6 for gene set analyses.

#### Specificity scores

To assess the specificity of expression for each gene across development, we calculated specificity scores in PsychENCODE data, using a similar specificity metric as Skene and colleagues^97^. First, we employed a normalization procedure that scaled the expression of a gene in each developmental window based on its relative expression in other windows, using the developmental periods defined by PsychENCODE investigators^98^. This was achieved by computing the average expression of each gene within each window as 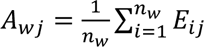, where *A_wj_* denotes the average expression of gene *j* for window *w*, *E_ij_* represents the expression level of gene *j* in sample *i* from window *w*, and *n_w_* is the number of samples associated with window *w*. We then calculated the specificity score, *S*, for each gene in each window as 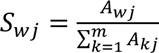, where *m* is the total number of windows.

### Genetic correlations with other complex traits

Genomic SEM v0.0.5, which is based on LD score regression^99,100^, was used to estimate genetic correlations between general impulsivity and other complex traits, as well as between the impulsivity facets and these traits. In the present paper, we selected traits from three domains for study: (i) imaging-derived measures of cortical structure, (ii) accelerometer-based measures of physical activity throughout the day, and (iii) psychiatric outcomes.

Genetic correlations were estimated adhering to best practices, using standard reference panels and parameter settings. All GWAS summary statistics were processed with the *munge* function of Genomic SEM, which retained only HapMap3 SNPs outside of the major histocompatibility complex regions with an allele frequency ≥ 0.01 for analysis. Within each family of tests, *P* values were adjusted for multiple comparisons via a Bonferroni correction. Note that all genetic correlations with the common factor were subsequently examined with *Q*_Trait_ tests, as previously described^33^. This test parallels *Q*_SNP_ and *Q*_Gene_, as it evaluates the degree to which genetic relationships between model indicators and an external trait plausibly operate via a common pathway, aligning with the magnitude of factor loadings.

### Polygenic score analyses

PRS-CS^101^ v2021.06.04 and PLINK^102^ v2.00 were used to calculate polygenic scores for the impulsivity facets and the common factor of impulsivity. Briefly, we first used PRS-CS to apply a continuous shrinkage prior to SNP effect estimates and infer posterior SNP weights for 875,517 common SNPs (minor allele frequency ≥ 0.01) that were present across the HapMap3 and 1000 Genomes Project Phase 3v5 datasets. European ancestry samples from the 1000 Genomes Project Phase 3v5 reference panel were used to model LD among variants. We then used PLINK to calculate polygenic scores for individuals in each sample by summing all included variants weighted by the inferred posterior effect size for the effect allele. The resulting polygenic scores were subsequently standardized to unit variance.

This process was used to calculate polygenic scores in two independent datasets: the Adolescent Brain and Cognitive Development Study (ABCD)^60,61^ and the Vanderbilt University Medical Center Biobank (BioVU)^62,63^. The former was used to examine associations between genetic liability for impulsivity and a select number of psychological outcomes in children, while the latter was used to examine associations between genetic liability for impulsivity and a wide array of medical outcomes, as described below.

#### Adolescent Brain and Cognitive Development Study

We used the ABCD Study (*n* = 4,314) to examine relationships between liability for impulsivity and closely related psychological outcomes in children. Specifically, we sought to characterize associations between impulsivity polygenic scores and eight outcomes measured at baseline: the five UPPS-P subscales (negative urgency, [lack of] premeditation, [lack of] perseverance, positive urgency, and sensation seeking)^40^ and three subscales from the Child Behavior Checklist (CBCL) (attention, externalizing, and internalizing problems)^64,65^.

To accomplish this, we used the lavaan R package (version 0.6-12) to conduct multiple regression analyses with DWLS estimation. For each outcome, we first fit a baseline model that only included sex, age, and the first 10 principal components of ancestry as covariates. We then fit models that iteratively added each polygenic score to the model, freely estimating the effect of the polygenic score on the outcome while adjusting for covariates. The variance explained by each score was calculated as Δ*R*^2^ = *R*^2^_full_ - *R*^2^_covariates_. As results revealed that the common factor polygenic score performed quite well in terms of Δ*R*^2^, we next sought to determine if the pattern of effects was consistent with a common pathway model. Below, we describe a novel means of testing for facet-specific relationships that is conceptually similar to *Q*_SNP_, *Q*_Gene_, and *Q*_Trait_ tests, but allows us to test whether individual effects deviate from expectation.

Recall that facet-level effects are expected to scale proportionally with the magnitude of unstandardized factor loadings if the common pathway model is valid. Thus, we began by establishing the expected beta coefficient for each facet-level polygenic score based on the effect of the common factor polygenic score and the facet’s unstandardized factor loading from the genomic factor model, such that

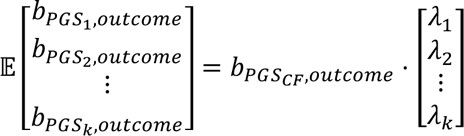

We then fit a follow-up model with a constraint imposed, wherein the effect of the polygenic score was fixed to its expected value. The freely estimated and constrained models were then compared using a likelihood ratio test, which allowed us to quantitatively determine whether deviations from expectation were statistically significant. Note that this approach is only suitable when the univariate GWAS (i.e., the input for Genomic SEM), have comparable statistical power. That is to say, if polygenic scores for the model indicators vary drastically in their predictive power, we may observe deviations that are not informative about the common pathway.

#### Vanderbilt University Medical Center Biobank

The genotyped BioVU sample (*n* = 66,915) served as the basis for examining associations between impulsivity polygenic scores and a broad range of medical phenotypes. The genotyping process and the quality control measures for this sample have been described in previous publications. Medical phenotypes, termed here as “phecodes”, were derived from the International Classification of Disease (ICD) diagnostic codes present in the electronic health records of participants. For a participant to be recognized as a case for a given phecode, we required that two separate ICD diagnostic code instances be present in their record. Only phecodes with at least 100 cases were considered for analysis. This resulted in 1,380 phecodes being included in the present study.

The PheWAS R package (version 0.99.5-2) was then used to conduct phenome-wide association analyses, where, for each of the phecodes, a logistic regression model was used to estimate the odds of each diagnosis based on each impulsivity polygenic score. Covariates included sex, median age from the longitudinal electronic health record data, and the first 10 principal components of ancestry. Phenome-wide significance was set at a Bonferroni-adjusted threshold of *P* ≤ 3.62e-5.

## Data Availability

No new data were collected for this study. Qualified researchers may download the impulsivity GWAS summary statistics through 23andMe, Inc. via a data use agreement that protects the privacy of their participants. Please visit https://research.23andme.com/collaborate/#dataset-access/ for more information regarding data access.

## Acknowledgments

We would like to thank the research participants and employees of 23andMe for making this work possible. The following members of the 23andMe Research Team contributed to this study: Stella Aslibekyan, Adam Auton, Elizabeth Babalola, Robert K. Bell, Jessica Bielenberg, Jonathan Bowes, Katarzyna Bryc, Ninad S. Chaudhary, Daniella Coker, Sayantan Das, Emily DelloRusso, Sarah L. Elson, Nicholas Eriksson, Teresa Filshtein, Pierre Fontanillas, Will Freyman, Zach Fuller, Chris German, Julie M. Granka, Karl Heilbron, Alejandro Hernandez, Barry Hicks, David A. Hinds, Ethan M. Jewett, Yunxuan Jiang, Katelyn Kukar, Alan Kwong, Yanyu Liang, Keng-Han Lin, Bianca A. Llamas, Matthew H. McIntyre, Steven J. Micheletti, Meghan E. Moreno, Priyanka Nandakumar, Dominique T. Nguyen, Jared O’Connell, Aaron A. Petrakovitz, G. David Poznik, Alexandra Reynoso, Shubham Saini, Morgan Schumacher, Leah Selcer, Anjali J. Shastri, Janie F. Shelton, Jingchunzi Shi, Suyash Shringarpure, Qiaojuan Jane Su, Susana A. Tat, Vinh Tran, Joyce Y. Tung, Xin Wang, Wei Wang, Catherine H. Weldon, Peter Wilton, Corinna D. Wong.

We would also like to add that the current analyses would not have been possible without the enormous efforts put forth by the investigators and participants from the Adolescent Brain Cognitive Development Study, BioVU, ENIGMA, iPSYCH, PsychENCODE, Psychiatric Genomics Consortium, and UK Biobank.

## Author Contributions

T.T.M. and S.S-R. conceived the study. The study protocol was developed by T.T.M. with input from S.S-R. T.T.M. was the lead analyst responsible for this study with assistance from J.D.T., M.J., and Y.Z. T.T.M. drafted the manuscript, with substantive contributions from J.W.S., A.A.P., and S.S-R. T.T.M. prepared the figures and tables with assistance from J.D.T., M.J., Y.Z., M.L.W., and C.M.W. Various aspects of study design and framing benefitted from advice and feedback from D.E.G., A.D.G., M.L.W., R.G.F., L.K.D., A.R., E.M.T-D., K.W.C., and T.G. All authors critically reviewed the manuscript and contributed to its final draft.

