## Supplementary Information and Figures for "The pleiotropic architecture of human impulsivity across biological scales"

| | |
|---|---|
| Word Count: | 881 (excluding figure legends and tables) |
| Tables: | 16 |
| Figures: | 20 |
| References: | 7 |

### Corresponding Author

| | |
|---|---|
| Name: | Travis T. Mallard, Ph.D. |
| Email Address: | |
| Mailing Address: | Psychiatric and Neurodevelopmental Genetics Unit, Center for Genomic Medicine, 185 Cambridge Street, Boston, MA 02114 |

**Investigating the factor structure of impulsivity facets**

We used linkage disequilibrium score regression[1,2] to estimate the genetic covariances among impulsivity facets in the present study. Generally, the BIS and UPPS-P phenotypes tended to show patterns of moderate-to-large genetic overlap with each other (mean $r_g = 0.434$) – with one exception. Sensation seeking was relatively weakly related to other traits (maximum $r_g = 0.337$ between sensation seeking and motor impulsivity). However, subsequent examination of the standardized genetic covariance matrix revealed that the first eigenvector explained 54% of the total genetic variance among impulsivity phenotypes. We calculated the tau-equivalent and congeneric reliability coefficients of the eight impulsivity facets as 0.853 and 0.869, respectively, which suggests that the traits are cohesively measuring the same underlying construct with good-to-excellent internal consistency.

Two non-graphical scree tests were used to determine the number of genomic factors needed to parsimoniously describe the data: the acceleration factor and optimal coordinates. Both were performed with the *nFactors* R package[3]. The acceleration factor is used to identify the point of inflection on the scree plot, which indicates that the subsequent factors explain relatively little variance, while the optimal coordinate determines the number of factors to retain based on comparisons of observed eigenvalues with their projected estimates. Here, the two analytic tests yielded different results, as the acceleration factor indicated only one factor was necessary while the optimal coordinates suggested that three factors might be more appropriate.

We proceeded to use these results to guide an exploratory factor analysis (EFA) of the data, specifying one- to three-factor solutions with promax rotation (i.e., allowing for the factors to be correlated with one another). In brief, the EFA results indicated that including additional factors did not yield a substantially more informative solution. A single factor explained 50% of the total SNP-based genetic variance across the study phenotypes, while two- and three-factor solutions only explained an additional 9% and 16%, respectively. Moreover, the one-factor solution was more parsimonious and interpretable than the alternatives, with indicators exhibiting strong and positive loadings on the common factor.

Based on these results, we proceeded to fit a single common factor model via confirmatory factor analysis (CFA) with Genomic Structural Equation Modeling[4]. Briefly, this model showed acceptable fit to the data ($\chi^2[20] = 608.958$, comparative fit index [CFI] = 0.899, standardized root mean squared residual [SRMR] = 0.103; **Figure S1**). Loadings on this common factor were generally quite strong (mean $\lambda$ = 0.666, median $\lambda$ = 0.785, average variance extracted [AVE] = 49.31%). However, examination of the residual covariance matrix suggested that there was complexity not accounted for by the common factor – largely between urgency traits and sensation seeking.

We calculated the residual covariance $Z$ statistics for the common factor model and found three significant residual covariances: negative urgency and positive urgency, negative urgency and sensation seeking, and sensation seeking and motor impulsivity. We subsequently expanded the base model, allowing these three pairs of residual variances to correlate. This revised model showed good fit to the data ($\chi^2[17] = 325.079$, CFI = 0.947, SRMR = 0.08; **Figure S1**), as it closely approximated the observed genetic covariance matrix. Loadings were very stable across these two specifications (mean $|\Delta\lambda| = 0.023$, median $|\Delta\lambda| = 0.016$).

Note that we tested alternative models with a greater number of factors, but they did not outperform the common factor approach. For example, guided by the EFA results, we fit a two correlated factors model where motor, nonplanning, perseverance, premeditation, and sensation seeking comprised one factor, while attentional, negative urgency, nonplanning, and positive urgency comprised the other factor. Notably, the correlation between these two factors was large ($r_g = 0.674$) and factor loadings were quite variable across F1 (mean $\lambda = 0.591$, median $\lambda = 0.478$) and F2 (mean $\lambda = 0.737$, median $\lambda = 0.815$). While the model showed a comparably good fit to the data ($\chi^2[18] = 398.154$, CFI = 0.935, SRMR = 0.078), interpretation is less clear given the highly correlated factors, variable factor loadings, and cross-loading indicator. Similarly, we found that a three correlated factors model showed good fit ($\chi^2[15] = 315.94$, CFI = 0.948, SRMR = 0.078), but suffered from the same issues without providing substantial improvements to explanatory power (two-factor model AVE = 57.4%, three-factor model AVE = 67.07%).

**Comparing the topography of cortical maps**

To better understand the spatial organization of genomic relationships between impulsivity and cortical structure, we analyzed the distribution of genetic effects using two canonical cortical atlases. Specifically, we characterized how $r_g$ and $Q_{\text{Trait}}$ estimates varied as a function of the cytoarchitectural classes defined by von Economo and Koskinas[5], as well as the functional intrinsic connectivity networks derived from resting-state functional magnetic resonance imaging by Yeo and Krienen[6]. We matched labels from these categorical parcellations to the corresponding Desikan–Killiany regions using a maximal overlap criterion.

For both atlases, we used a Kruskal-Wallis rank sum test to compute an omnibus $F$ statistic that quantified the degree to which $r_g$ and $Q_{\text{Trait}}$ estimates differed as a function of cortical classes or networks. Here, we used a "spin" test to evaluate statistical significance[7], which compared the observed test statistic against an empirical null distribution of 10,000 spatially permutated test statistics. Critically, this allowed us to compare neuroimaging maps while accounting for the spatial contiguity and hemispheric symmetry of the cortex.

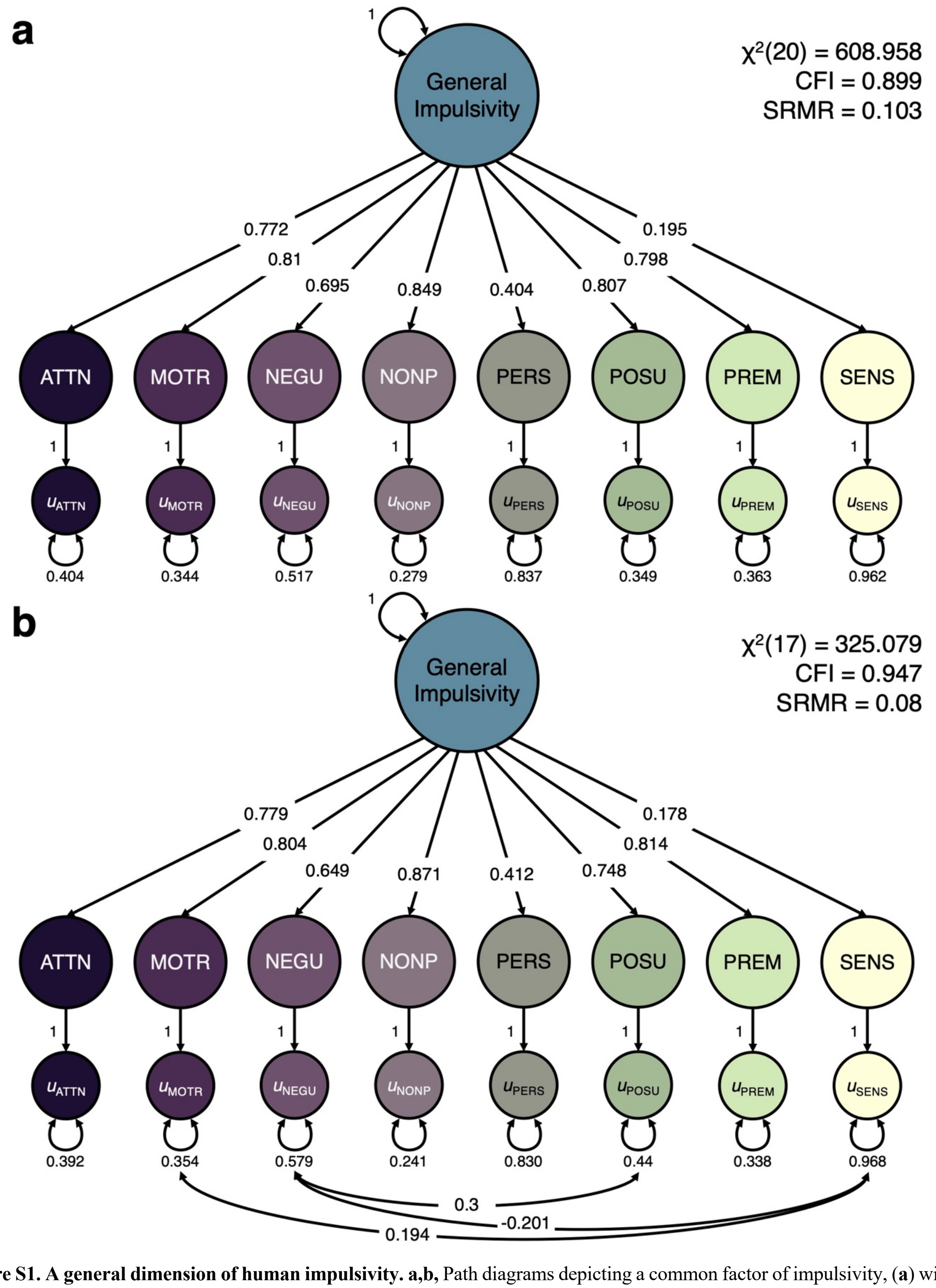


**Figure S1. A general dimension of human impulsivity. a,b,** Path diagrams depicting a common factor of impulsivity, (**a**) without and (**b**) with correlated residuals, where all facets of impulsivity load onto a single common factor. One-headed arrows pointing from independent variables to dependent variables reflect regression relationships. Two-headed arrows connecting variables represent covariance relationships. A two-headed arrow connecting a variable to itself denotes (residual) variance. Model fit statistics are inlaid within each panel. ATTN = attentional, MOTR = motor, NEGU = negative urgency, NONP = nonplanning, PERS = lack of perseverance, POSU = positive urgency, PREM = lack of premeditation, SENS = sensation seeking.

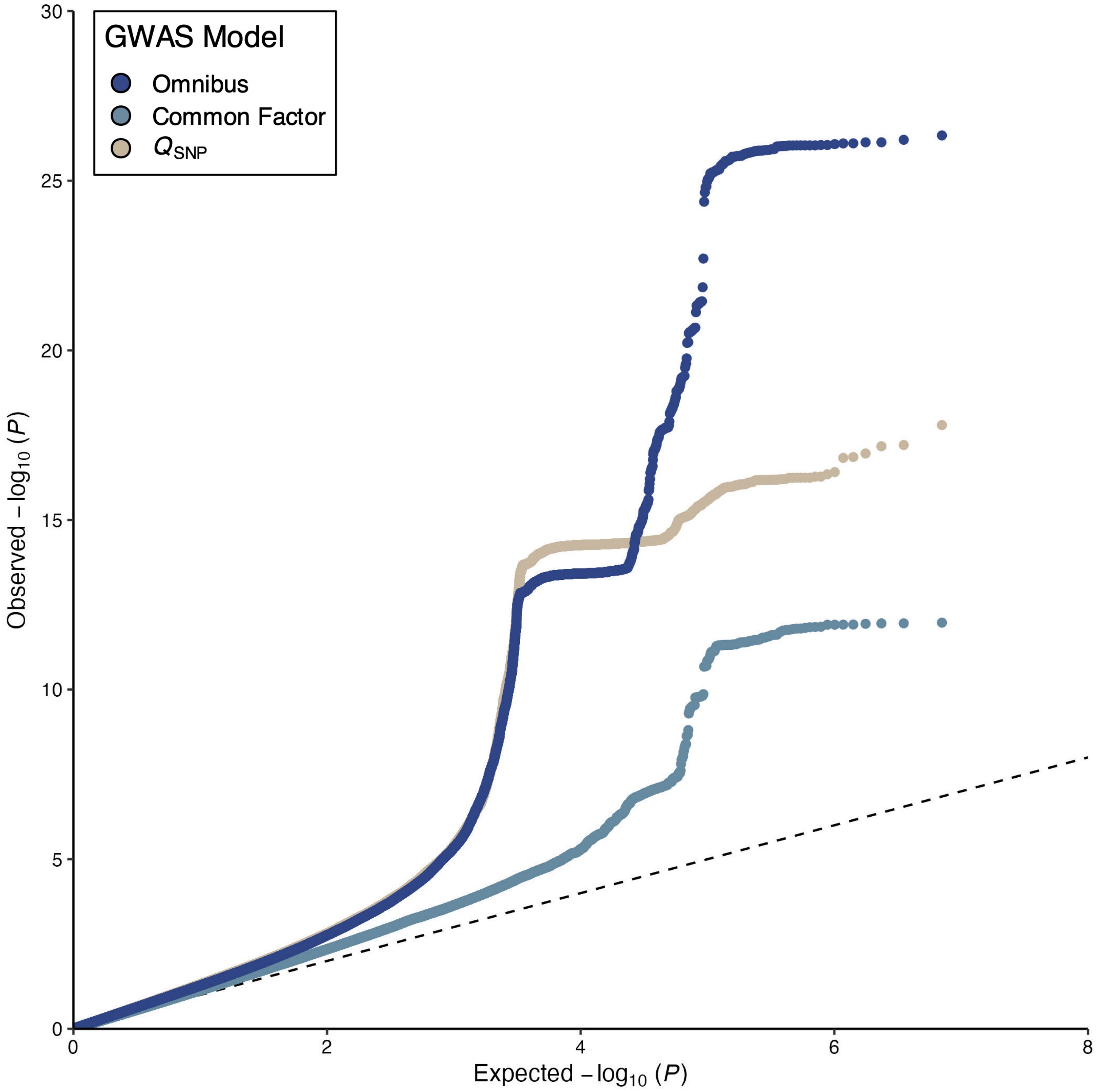


**Figure S2. Polygenic signal of three multivariate GWAS models.** Quantile-quantile plot for the multivariate GWAS results, illustrating stronger polygenic signal for both the omnibus and $Q_{SNP}$ results relative to the common factor model. The y-axis corresponds to the observed distribution of *P* values, while the x-axis corresponds to the expected distribution of P under the null. The theoretical null is plotted as a dashed black line.

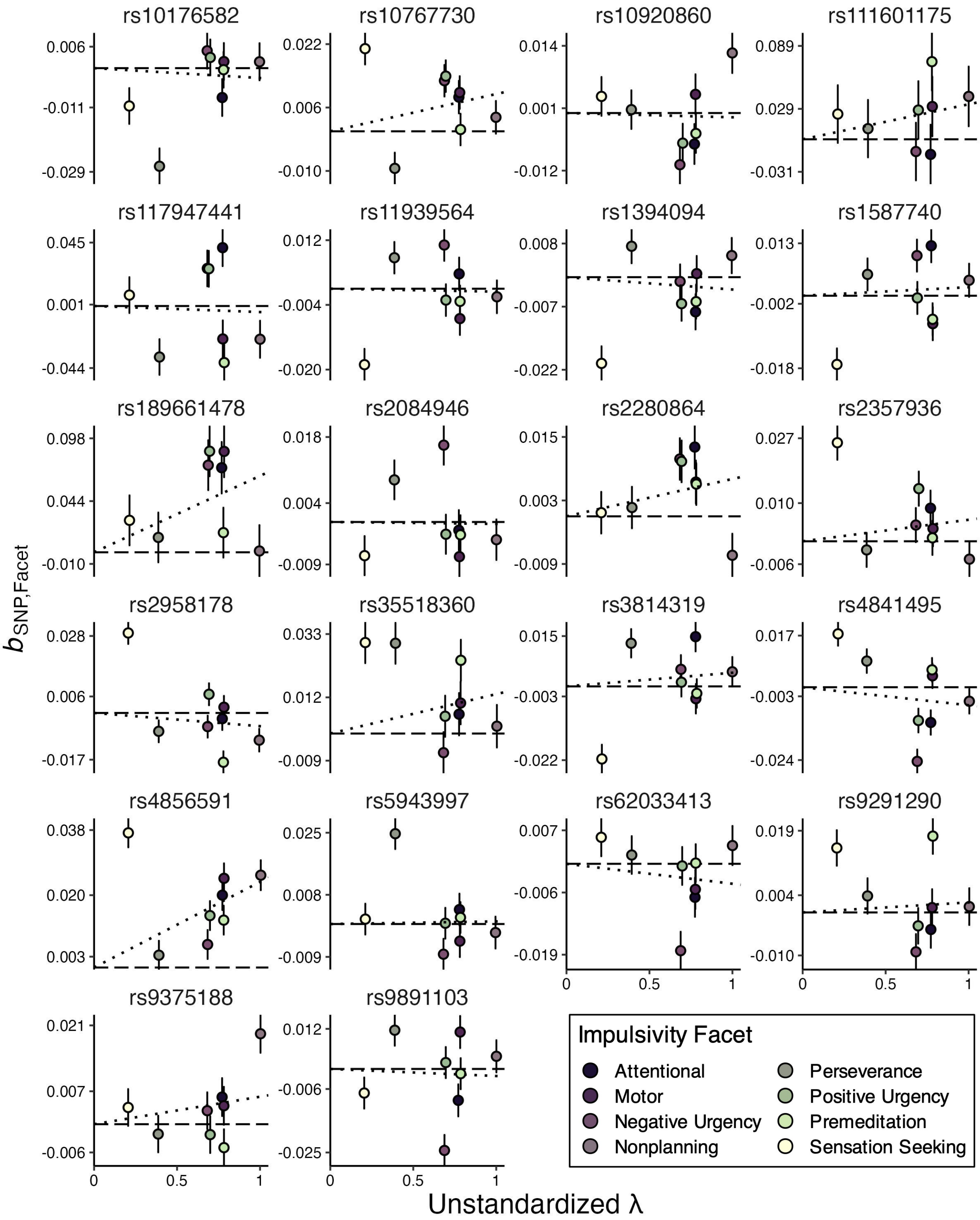


**Figure S3. Univariate SNP effects as a function of factor loadings for all significant $Q_{SNP}$ loci.** Scatter plots depicting the indicator GWAS estimates for all independent $Q_{SNP}$ loci as a function of unstandardized factor loadings. Error bars represent standard errors of the beta coefficient. The dotted black line represents the line of best fit, as estimated with linear regression with the intercept fixed to zero. Deviations from this line lend insight into how significant $Q_{SNP}$ $P$ values arise. For example, the larger negative effect of rs10176582 on (lack of) perseverance is inconsistent with the common factor model, driving the significant $Q_{SNP}$ $P$ value.

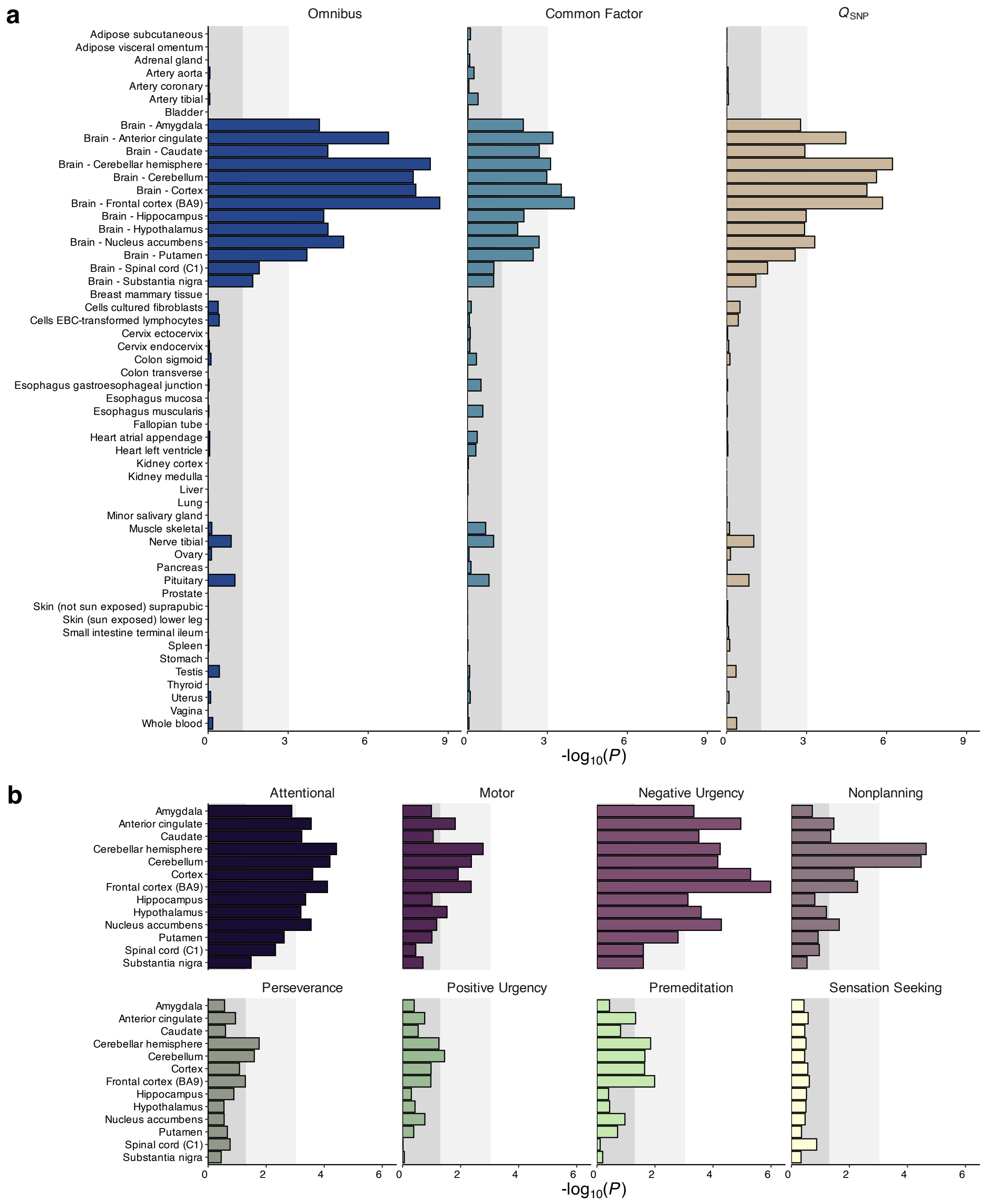


**Figure S4. Links between the genetic architecture of impulsivity and tissue-specific expression. a,b,** Bar charts of the gene property analysis for the (**a**) multivariate and (**b**) univariate GWAS results, illustrating patterns of enrichment for genomic signals in tissue-specific genes across the lifespan. Tissues measured in the Genotype-Tissue Expression (GTEx) v8 dataset are plotted on the y-axis, while the *P* value of enrichment is plotted on the x-axis on the $-\log_{10}$ scale. The dark gray band denotes the non-significant range of *P* values, the light gray bands reflect nominal significance, and white reflects significance at a Bonferroni-corrected threshold. For illustration purposes, only brain tissues are shown for the univariate results, as all non-brain tissues were nonsignificant.

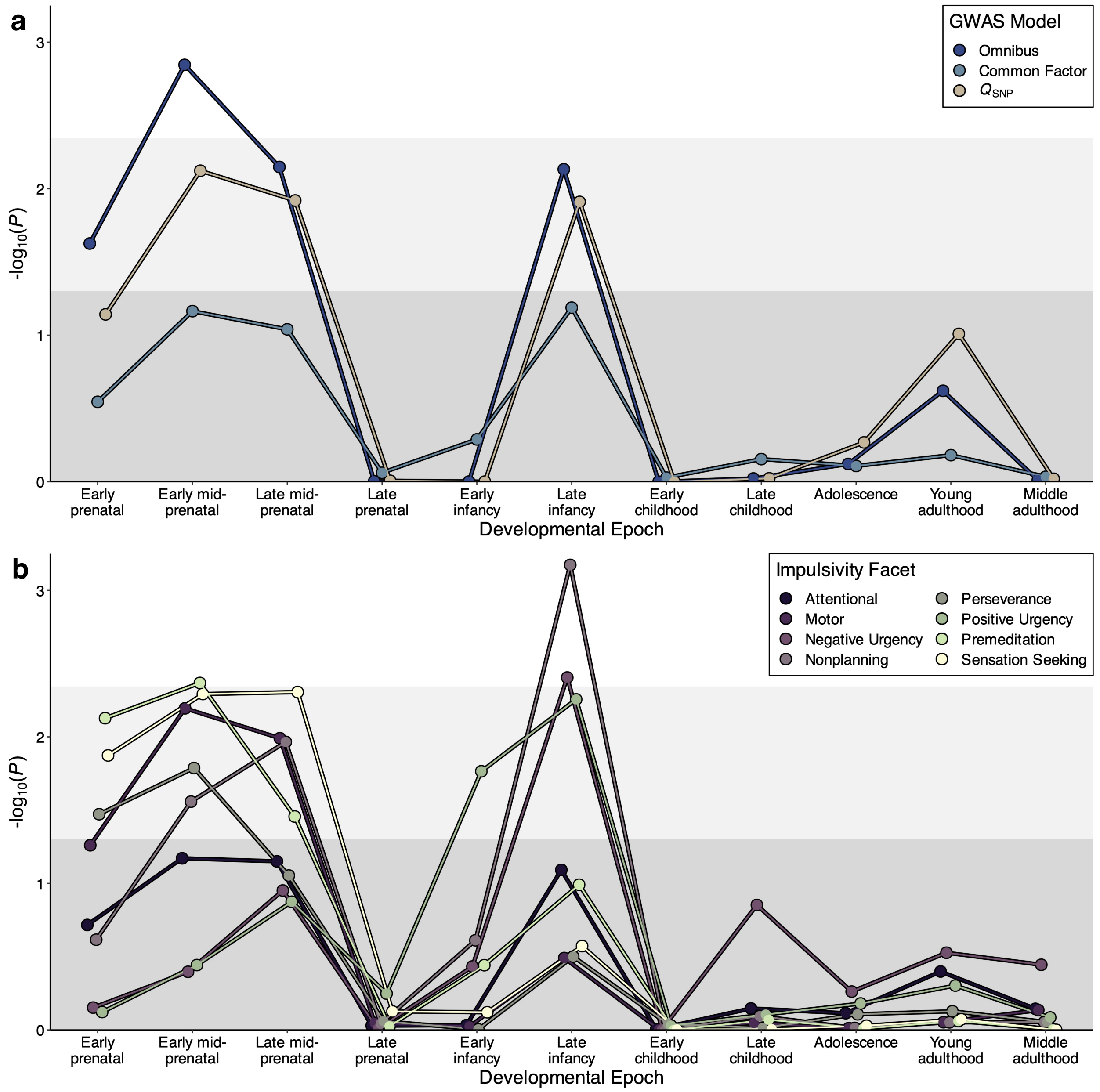


**Figure S5. Links between the genetic architecture of impulsivity and neurodevelopmental expression. a,b,** Line plots of the gene property analysis for the (**a**) multivariate and (**b**) univariate GWAS results, illustrating patterns of enrichment for genomic signals in developmentally specific genes across the lifespan. Developmental epochs measured in BrainSpan are plotted on the x-axis, while the *P* value of enrichment is plotted on the y-axis on the -$\log_{10}$ scale. The dark gray band denotes the non-significant range of *P* values, the light gray bands reflect nominal significance, and white reflects significance at a Bonferroni-corrected threshold.

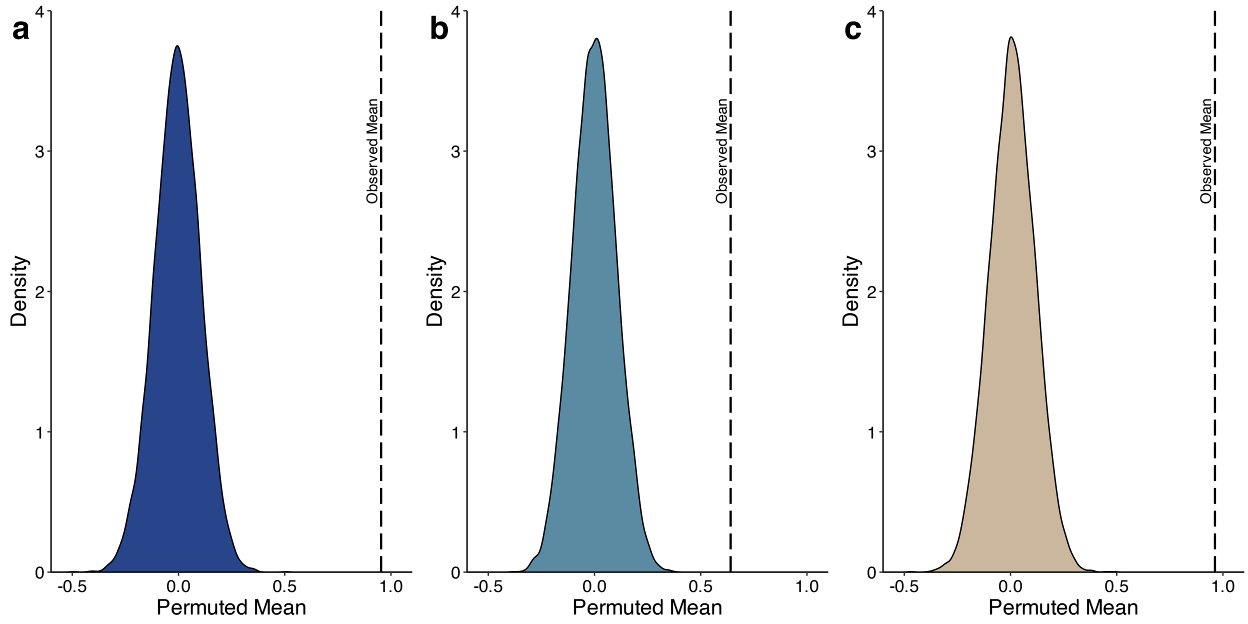


**Figure S6. Enrichment of synaptic biology gene sets in the genetic architecture of impulsivity. a,b,c,** Density plots illustrating the difference between the observed mean *Z* statistic of SynGO gene sets and the distribution of permuted mean *Z* statistics from equivalently sized comparison sets for the (**a**) omnibus, (**b**) common factor, and (**c**) $Q_{\mathrm{SNP}}$ GWAS results.

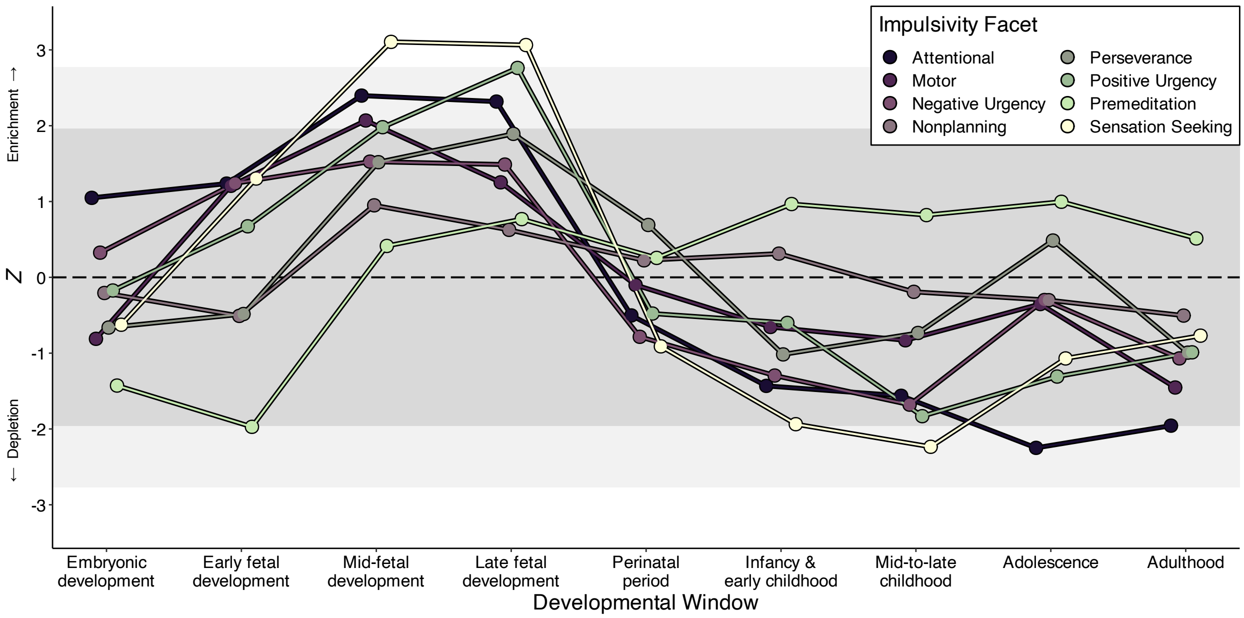


**Figure S7. Links between transcriptomic architecture and neurodevelopmental expression among impulsivity facets.** Line plot of gene property analysis for the facet-level TWAS results, illustrating patterns of enrichment and depletion of transcriptomic signals in developmentally specific genes across the lifespan. Developmental windows measured in PsychENCODE are plotted on the x-axis, while the $Z$ statistic of enrichment or depletion is plotted on the y-axis. The dark gray band denotes the non-significant range of $Z$ statistics, the light gray bands reflect nominal significance, and white reflects significance at a Bonferroni-corrected threshold.

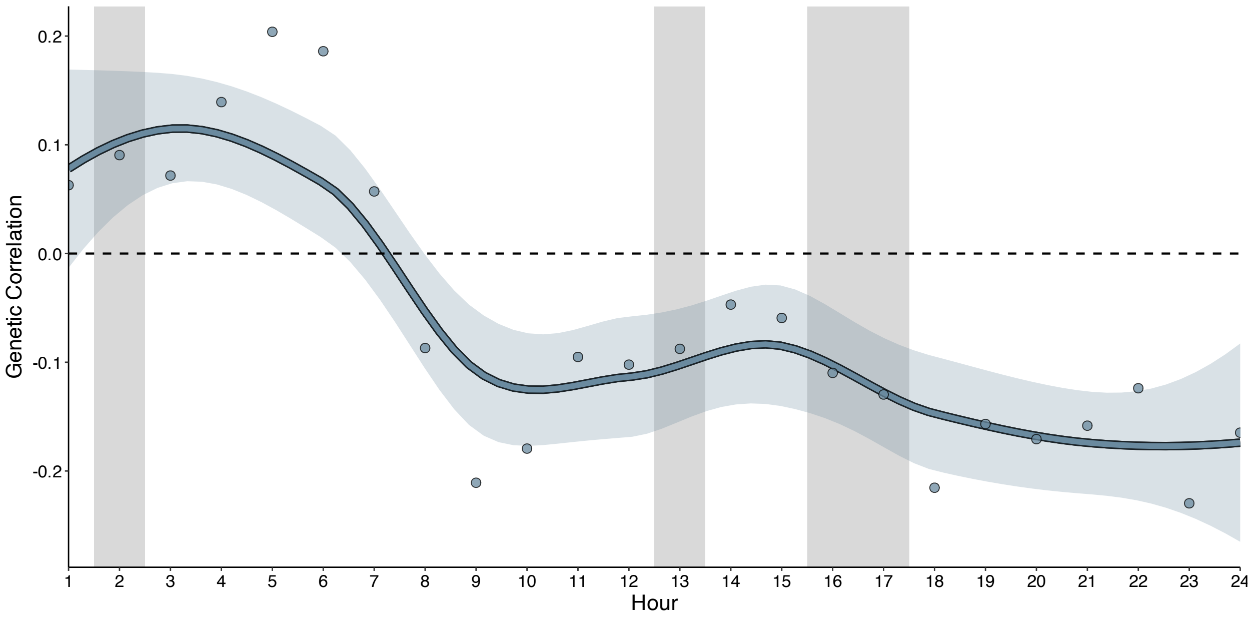


**Figure S8. Genetic relationships between the common factor of impulsivity and physical activity throughout the day.** Line plot of genetic correlations between the common factor of impulsivity and accelerometer-based measures of physical activity, as estimated with LOESS (with accompanying 95% confidence interval). The vertical gray bands highlight time periods when $Q_{\text{Trait}}$ *P* was non-significant (i.e., periods when the observed pattern of effects was consistent with a common pathway).

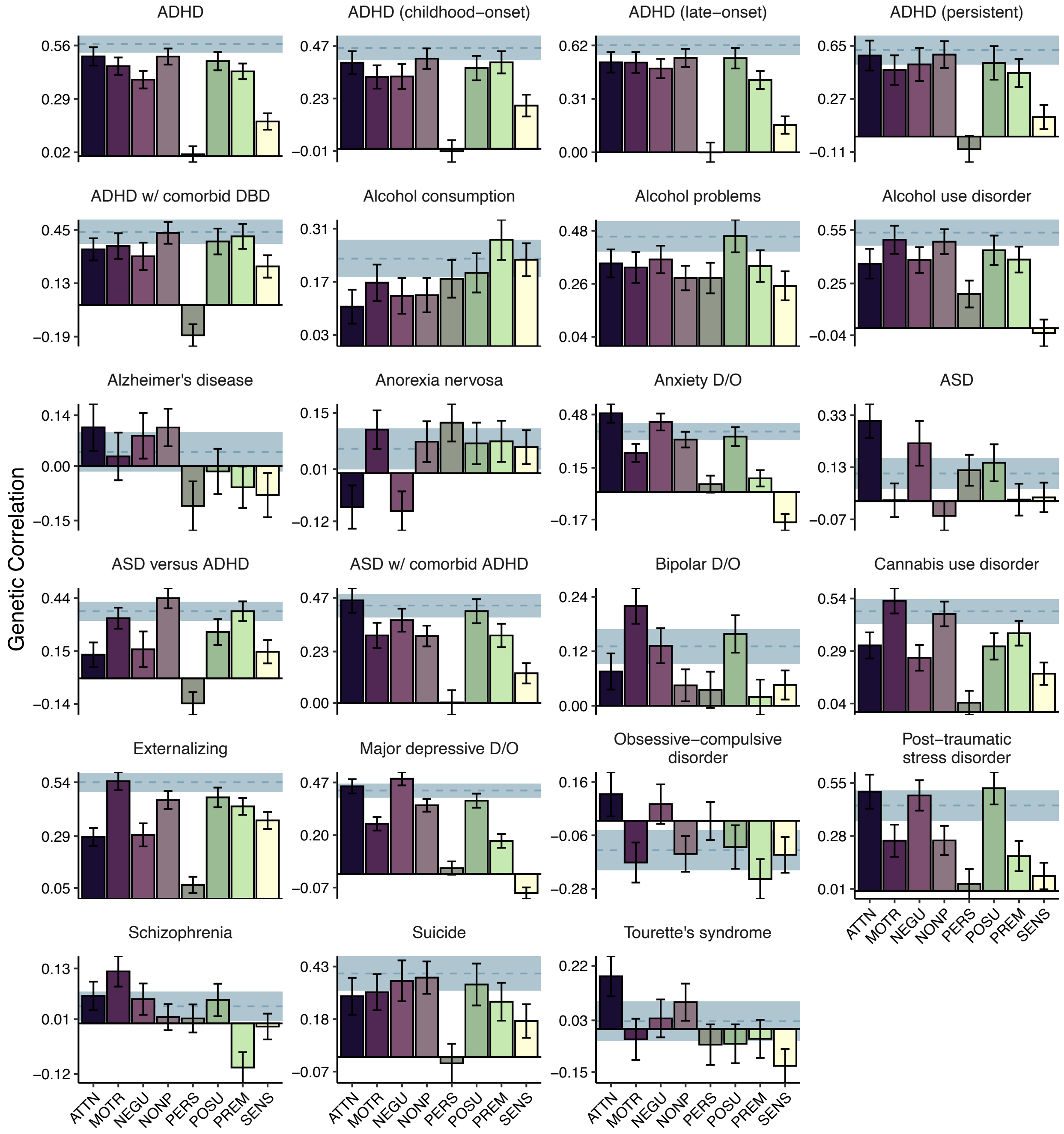


**Figure S9. Bivariate relationships between facets of impulsivity and psychiatric outcomes.** Bar charts depicting the genetic correlations between impulsivity facets and all tested psychiatric outcomes. Horizontal blue lines index the genetic correlation between an outcome and the impulsivity factor (and the corresponding standard error). ADHD = attention-deficit/hyperactivity disorder, ASD = autism spectrum disorder, DBD = disruptive behavioral disorder, D/O = disorder.

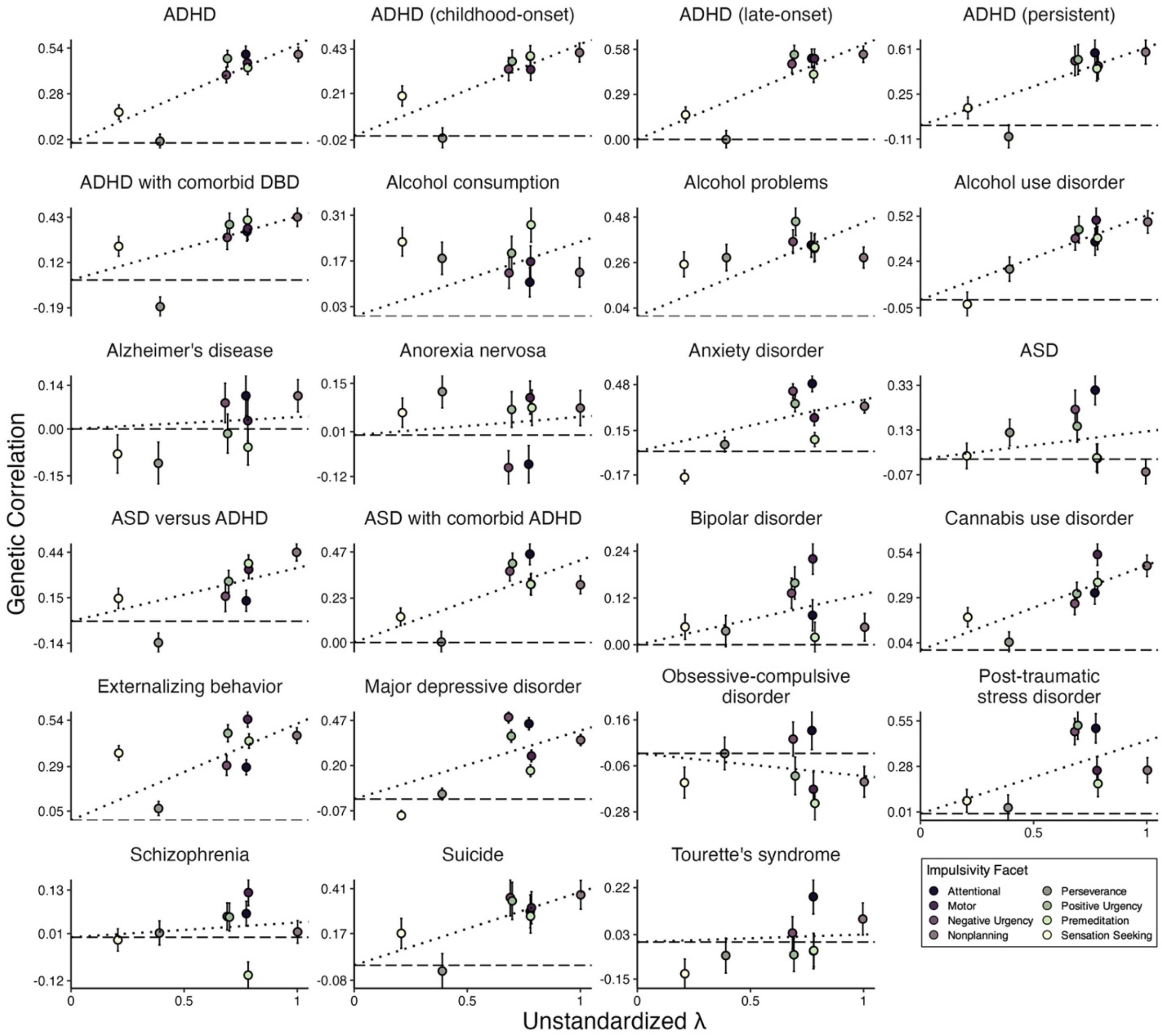


**Figure S10. Bivariate relationships between impulsivity facets and psychopathology as a function of factor loadings.** Scatter plots depicting the genetic correlations between impulsivity facets and all tested psychiatric outcomes as a function of unstandardized factor loadings. Error bars represent standard errors of the genetic correlation. The dashed line corresponds to a genetic correlation estimate of 0. The dotted black line represents the line of best fit, as estimated with linear regression with the intercept fixed to zero. Deviations from this line lend insight into how significant $Q_{\text{Trait}}$ $P$ values arise. For example, the null genetic correlation between ADHD and (lack of) perseverance is inconsistent with the common factor model, driving the significant $Q_{\text{Trait}}$ $P$ value. ADHD = attention-deficit/hyperactivity disorder, ASD = autism spectrum disorder, DBD = disruptive behavioral disorder.

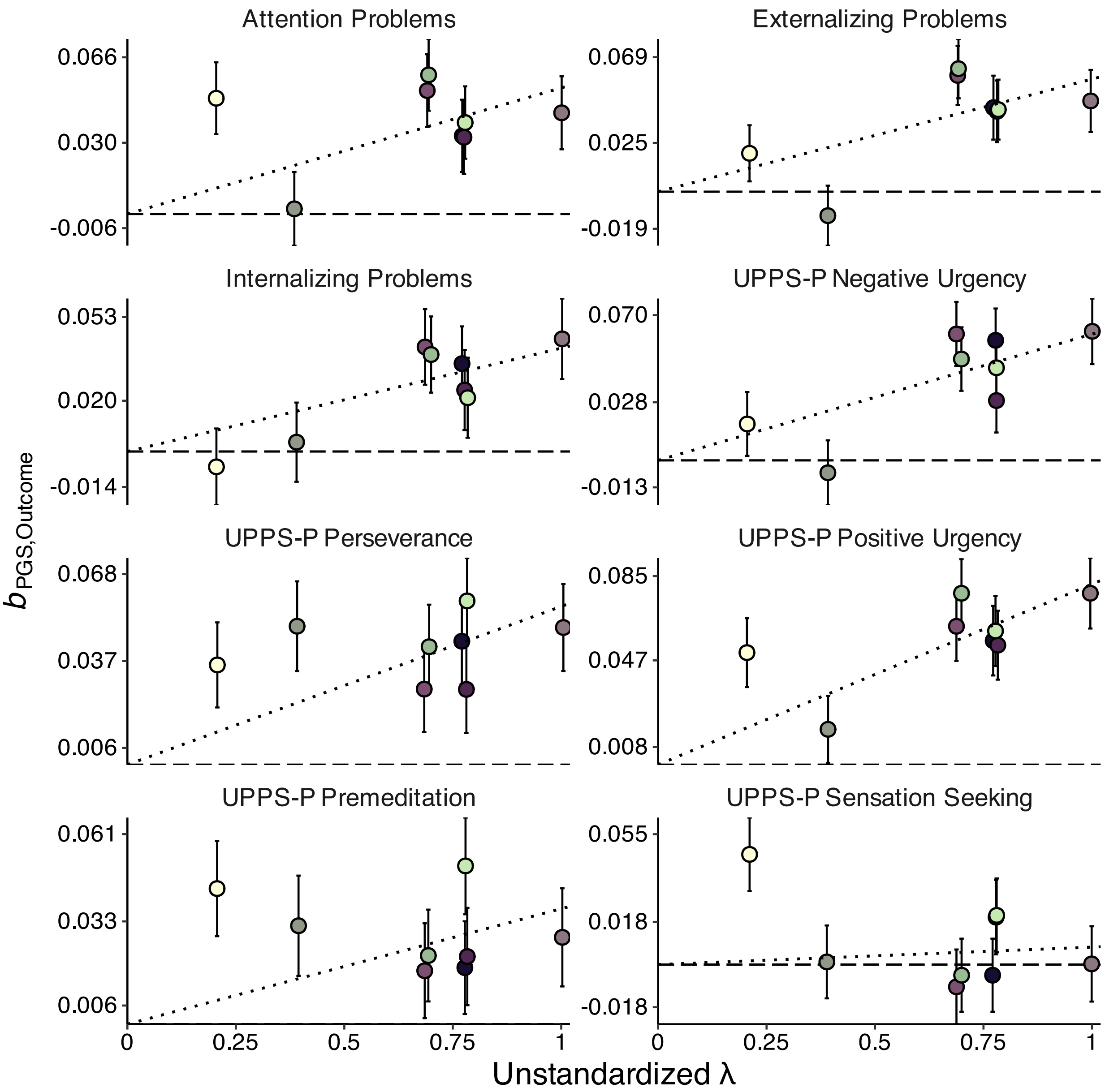


**Figure S11. Validity of common factor polygenic score for outcomes related to impulsivity in the ABCD Study.** Scatterplots depicting the indicator polygenic score effects as a function of unstandardized factor loadings in the Adolescent Brain Cognitive Development (ABCD) Study. Error bars represent the standard errors of the beta coefficient. The dotted black line represents the line of best fit, as estimated with linear regression with the intercept fixed to zero.

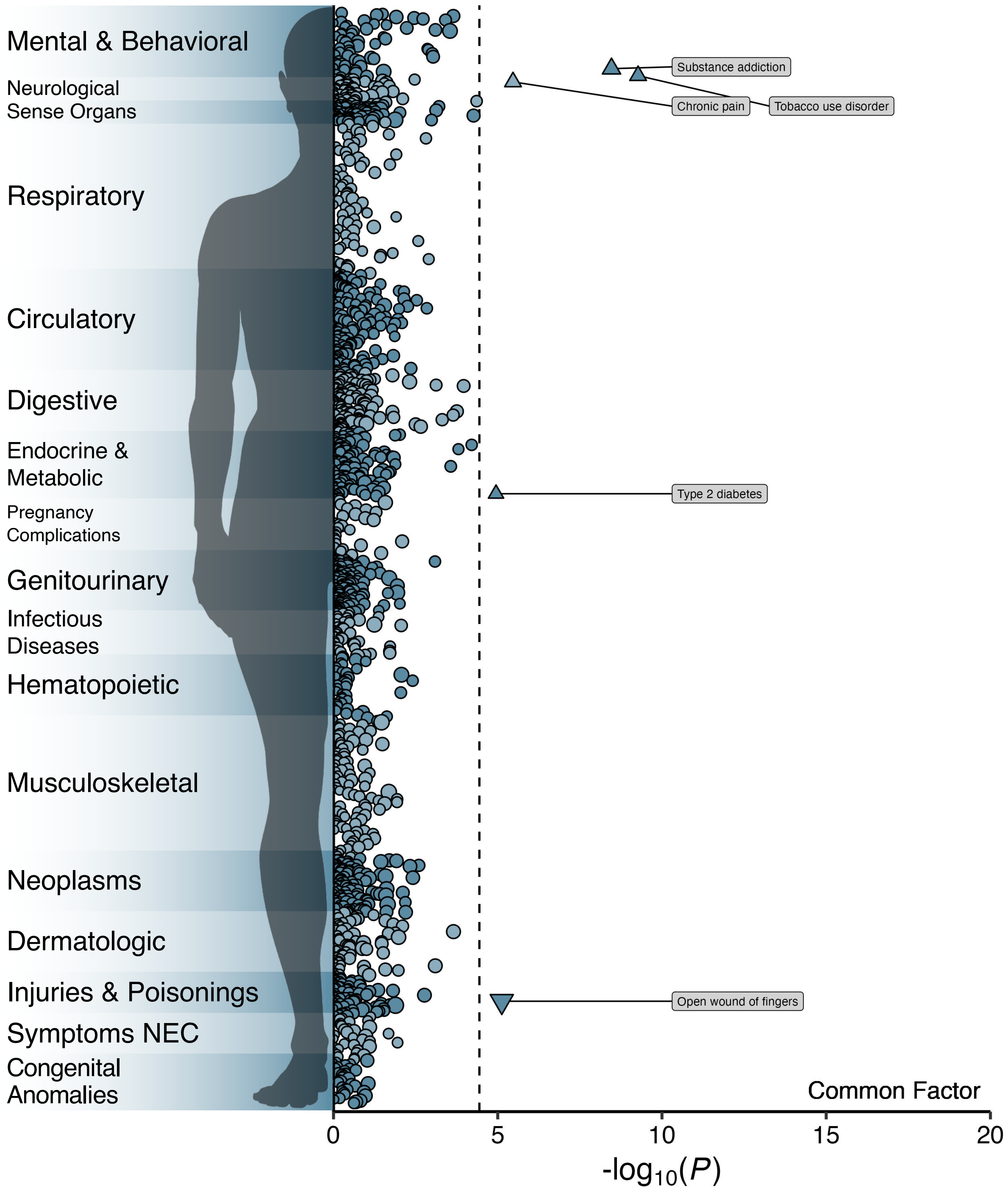


**Figure S12. Results of phenome-wide association study of common factor polygenic score in BioVU.** Manhattan plot of the associations between the common factor polygenic score and medical outcomes in BioVU. The y-axis refers to the category of medical outcome (or "phecode"), the x-axis refers to the statistical significance of the association on $-\log_{10}$ scale, and the dashed line denotes the Bonferroni-adjusted significance threshold. Circles denote associations that are nonsignificant after correction, upward-facing triangles are significant positive associations, and downward-facing triangles are significant negative associations. Size reflects the effect size, with larger points reflecting a larger absolute effect. For plotting purposes, some labels have been shortened or omitted. NEC = not elsewhere classified.

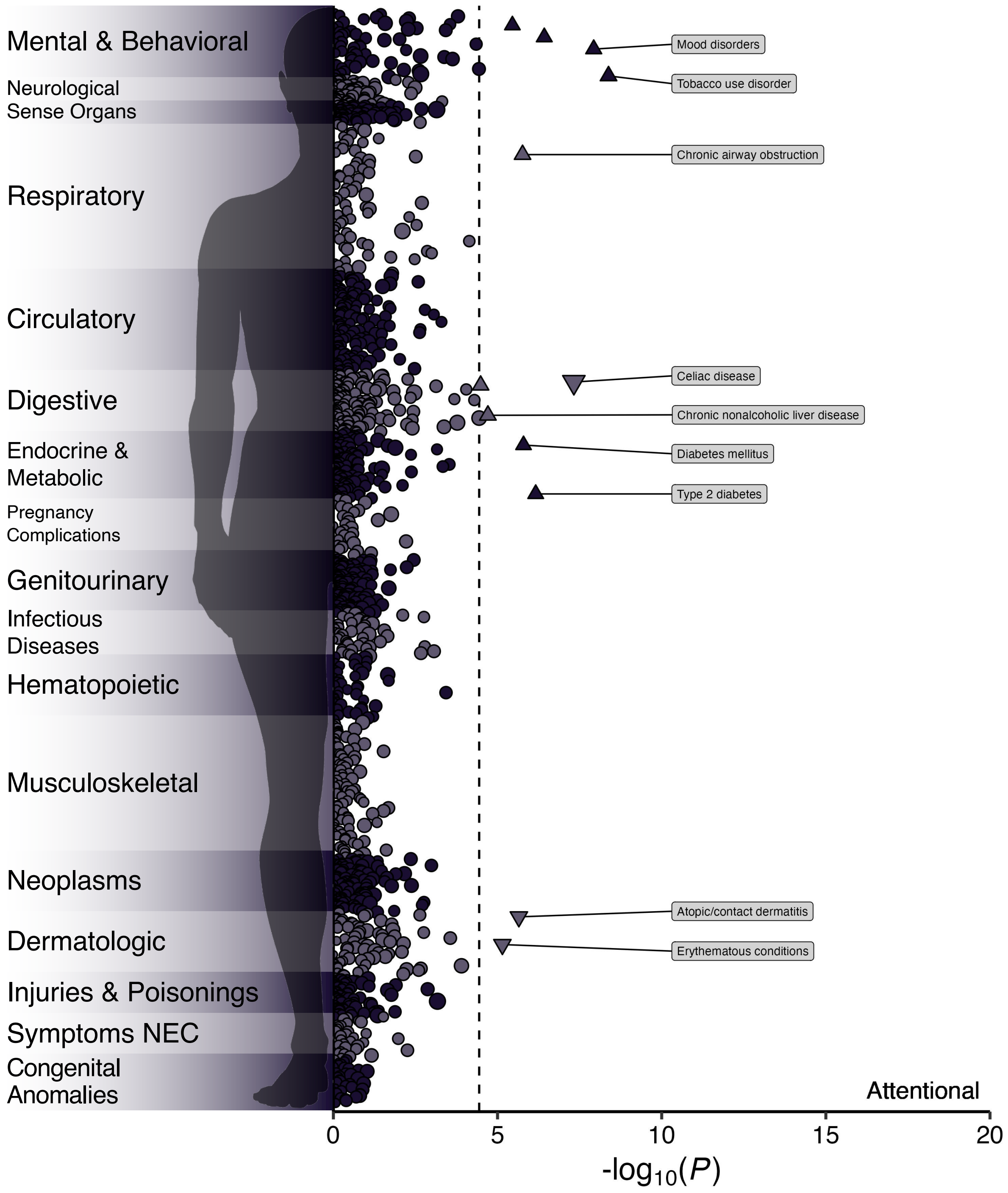


**Figure S13. Results of phenome-wide association study of attentional impulsivity polygenic score in BioVU.** Manhattan plot of the associations between the attentional impulsivity polygenic score and medical outcomes in BioVU. The y-axis refers to the category of medical outcome (or "phecode"), the x-axis refers to the statistical significance of the association on -$\log_{10}$ scale, and the dashed line denotes the Bonferroni-adjusted significance threshold. Circles denote associations that are nonsignificant after correction, upward-facing triangles are significant positive associations, and downward-facing triangles are significant negative associations. Size reflects the effect size, with larger points reflecting a larger absolute effect. For plotting purposes, some labels have been shortened or omitted. NEC = not elsewhere classified.

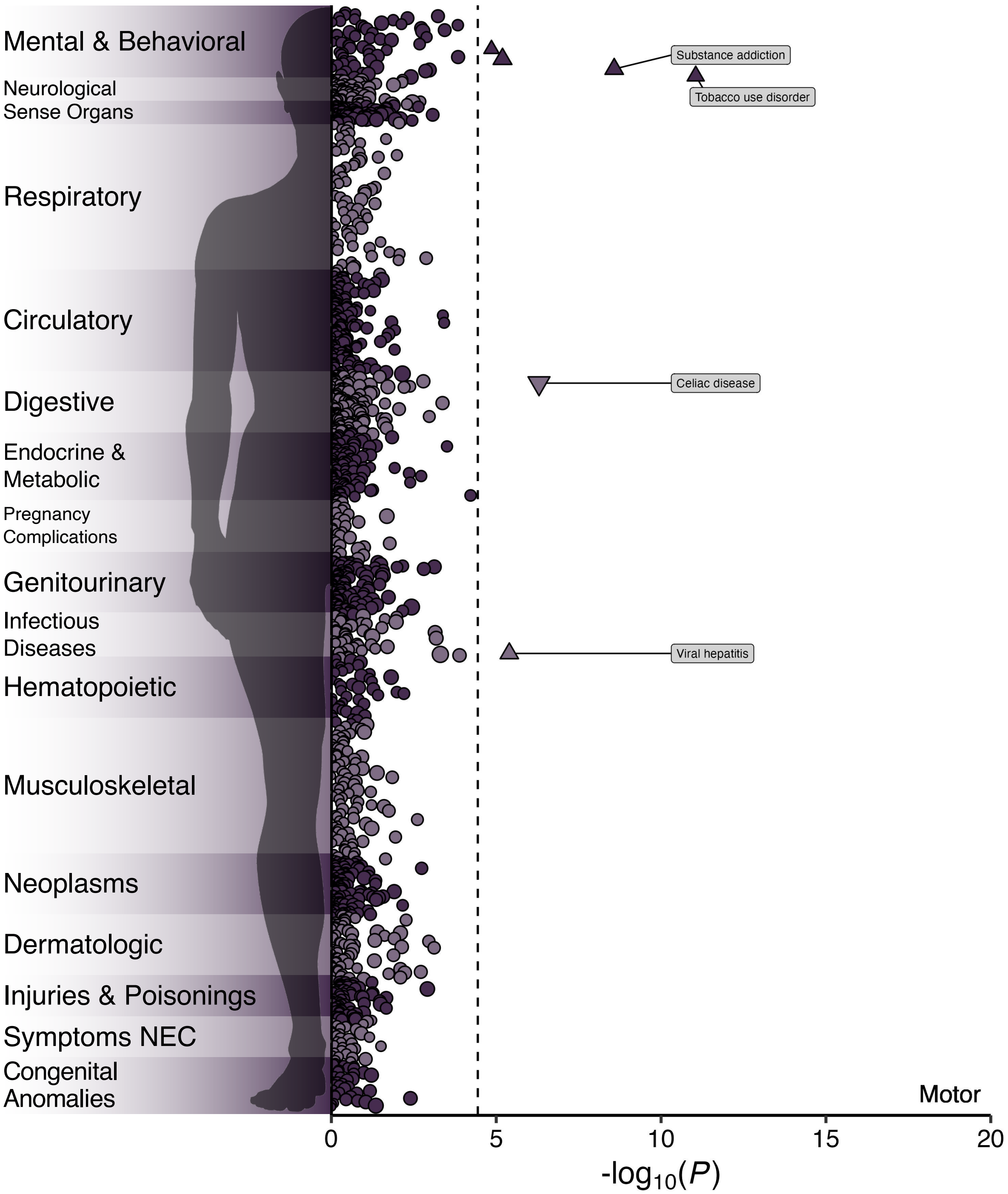


**Figure S14. Results of phenome-wide association study of motor impulsivity polygenic score in BioVU.** Manhattan plot of the associations between the motor impulsivity polygenic score and medical outcomes in BioVU. The y-axis refers to the category of medical outcome (or "phecode"), the x-axis refers to the statistical significance of the association on $-\log_{10}$ scale, and the dashed line denotes the Bonferroni-adjusted significance threshold. Circles denote associations that are nonsignificant after correction, upward-facing triangles are significant positive associations, and downward-facing triangles are significant negative associations. Size reflects the effect size, with larger points reflecting a larger absolute effect. For plotting purposes, some labels have been shortened or omitted. NEC = not elsewhere classified.

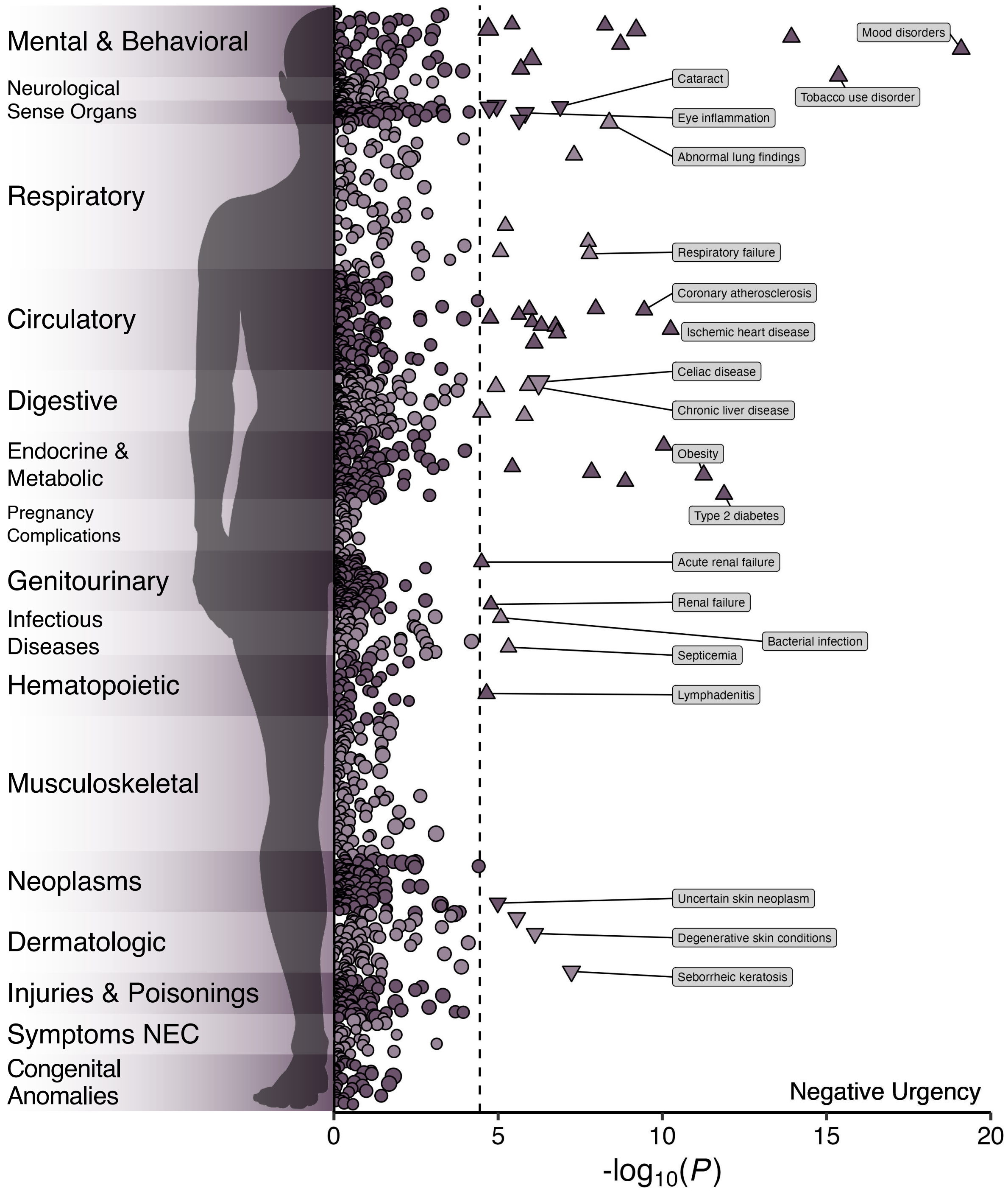


**Figure S15. Results of phenome-wide association study of negative urgency polygenic score in BioVU.** Manhattan plot of the associations between the attentional negative urgency polygenic score and medical outcomes in BioVU. The y-axis refers to the category of medical outcome (or "phecode"), the x-axis refers to the statistical significance of the association on $-\log_{10}$ scale, and the dashed line denotes the Bonferroni-adjusted significance threshold. Circles denote associations that are nonsignificant after correction, upward-facing triangles are significant positive associations, and downward-facing triangles are significant negative associations. Size reflects the effect size, with larger points reflecting a larger absolute effect. For plotting purposes, some labels have been shortened or omitted. NEC = not elsewhere classified.

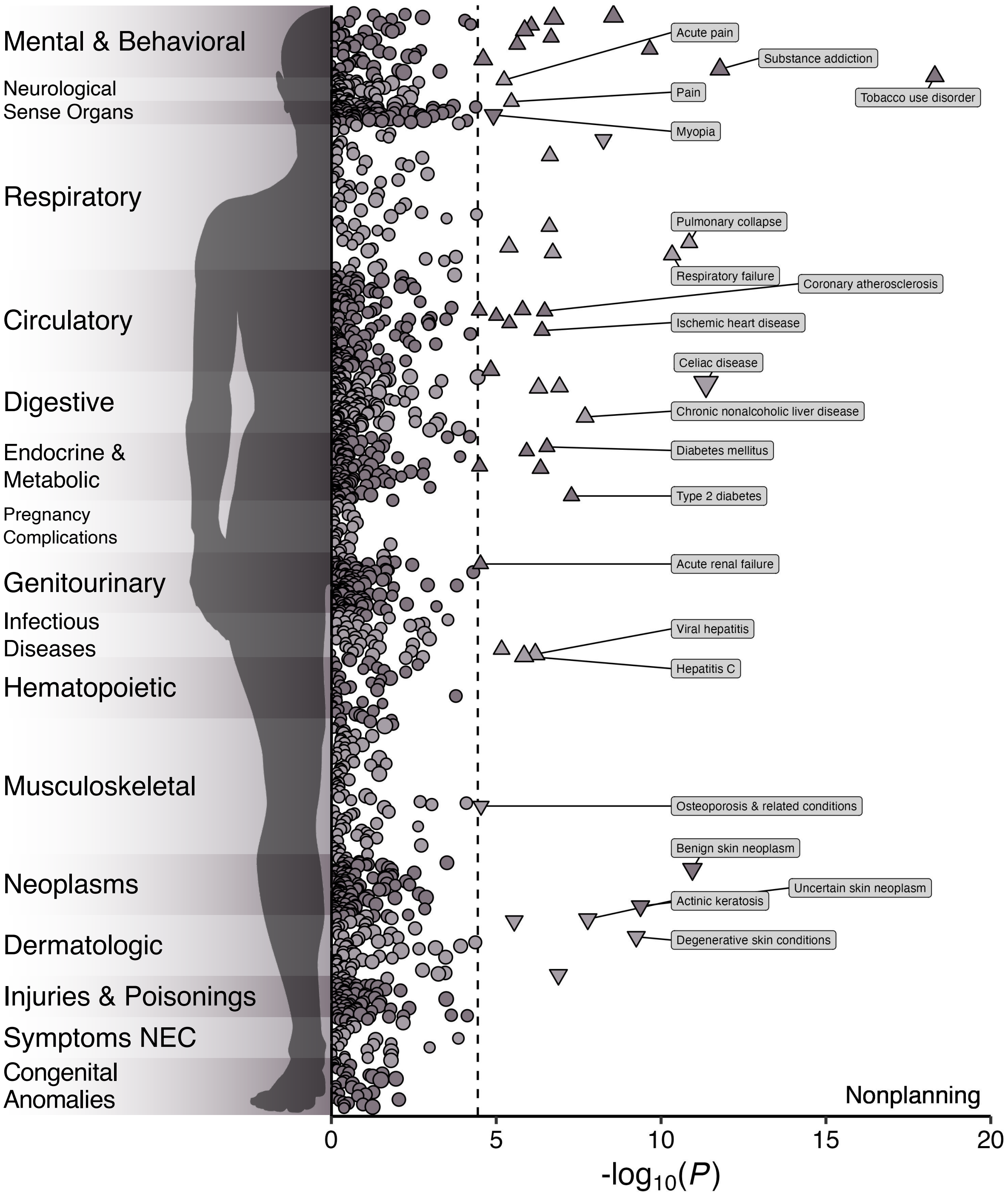


**Figure S16. Results of phenome-wide association study of nonplanning impulsivity polygenic score in BioVU.** Manhattan plot of the associations between the nonplanning impulsivity polygenic score and medical outcomes in BioVU. The y-axis refers to the category of medical outcome (or "phecode"), the x-axis refers to the statistical significance of the association on $-\log_{10}$ scale, and the dashed line denotes the Bonferroni-adjusted significance threshold. Circles denote associations that are nonsignificant after correction, upward-facing triangles are significant positive associations, and downward-facing triangles are significant negative associations. Size reflects the effect size, with larger points reflecting a larger absolute effect. For plotting purposes, some labels have been shortened or omitted. NEC = not elsewhere classified.

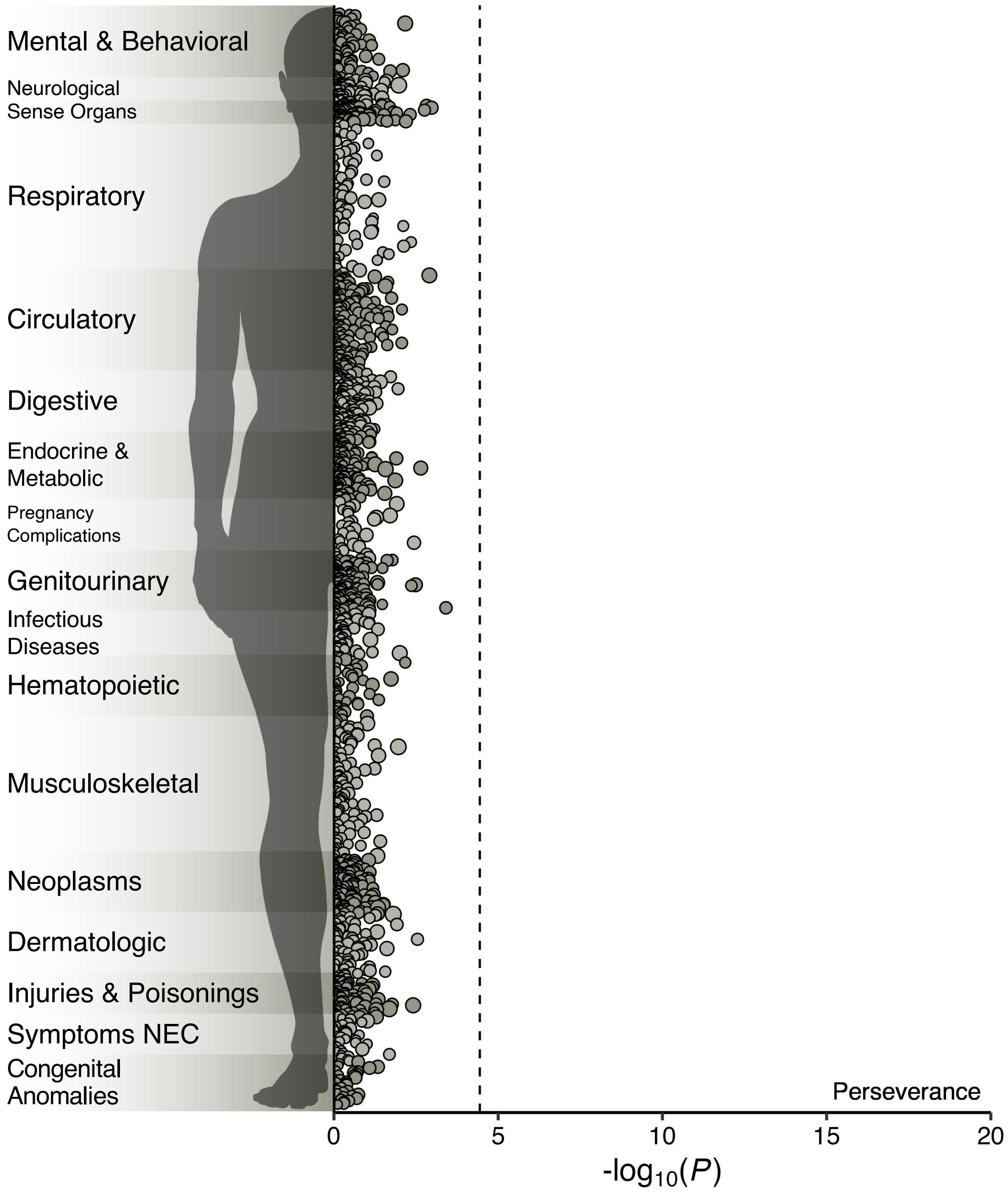


**Figure S17. Results of phenome-wide association study of (lack of) perseverance polygenic score in BioVU.** Manhattan plot of the associations between the (lack of) perseverance polygenic score and medical outcomes in BioVU. The y-axis refers to the category of medical outcome (or "phecode"), the x-axis refers to the statistical significance of the association on -log$_{10}$ scale, and the dashed line denotes the Bonferroni-adjusted significance threshold. Circles denote associations that are nonsignificant after correction, upward-facing triangles are significant positive associations, and downward-facing triangles are significant negative associations. Size reflects the effect size, with larger points reflecting a larger absolute effect. For plotting purposes, some labels have been shortened or omitted. NEC = not elsewhere classified.

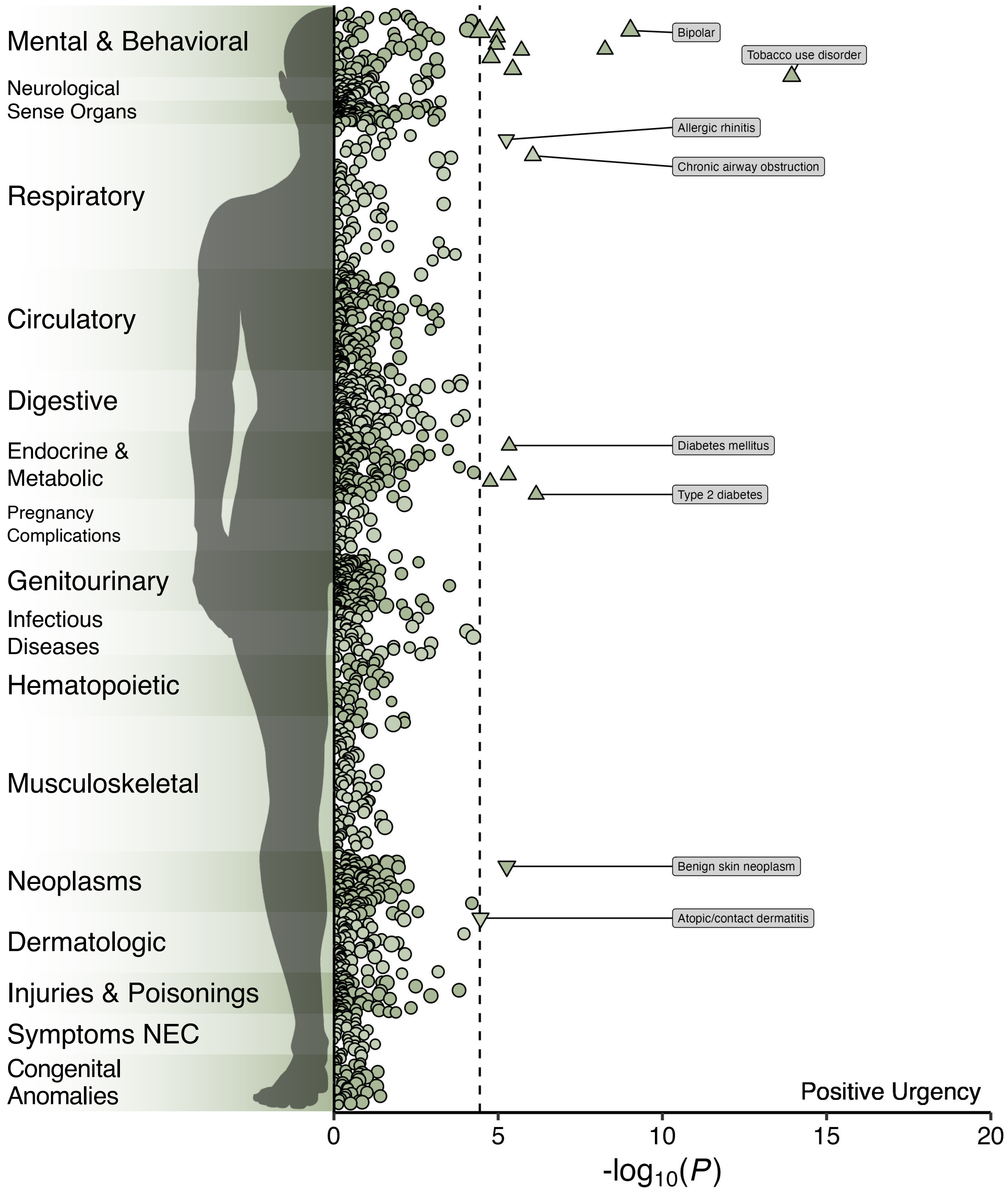


**Figure S18. Results of phenome-wide association study of positive urgency polygenic score in BioVU.** Manhattan plot of the associations between the positive urgency polygenic score and medical outcomes in BioVU. The y-axis refers to the category of medical outcome (or "phecode"), the x-axis refers to the statistical significance of the association on $-\log_{10}$ scale, and the dashed line denotes the Bonferroni-adjusted significance threshold. Circles denote associations that are nonsignificant after correction, upward-facing triangles are significant positive associations, and downward-facing triangles are significant negative associations. Size reflects the effect size, with larger points reflecting a larger absolute effect. For plotting purposes, some labels have been shortened or omitted. NEC = not elsewhere classified.

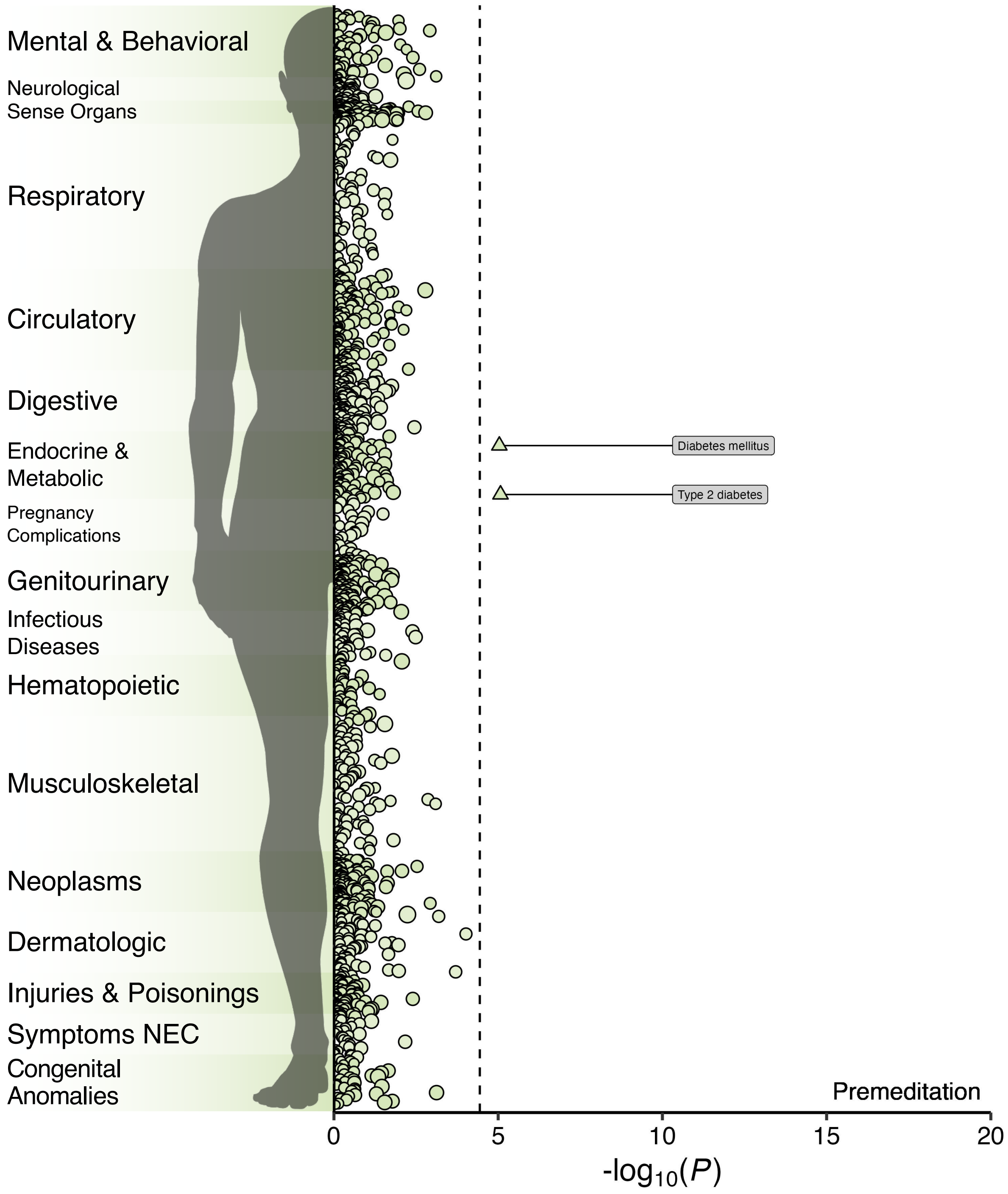


**Figure S19. Results of phenome-wide association study of (lack of) premeditation polygenic score in BioVU.** Manhattan plot of the associations between the (lack of) premeditation polygenic score and medical outcomes in BioVU. The y-axis refers to the category of medical outcome (or "phecode"), the x-axis refers to the statistical significance of the association on -$\log_{10}$ scale, and the dashed line denotes the Bonferroni-adjusted significance threshold. Circles denote associations that are nonsignificant after correction, upward-facing triangles are significant positive associations, and downward-facing triangles are significant negative associations. Size reflects the effect size, with larger points reflecting a larger absolute effect. For plotting purposes, some labels have been shortened or omitted. NEC = not elsewhere classified.

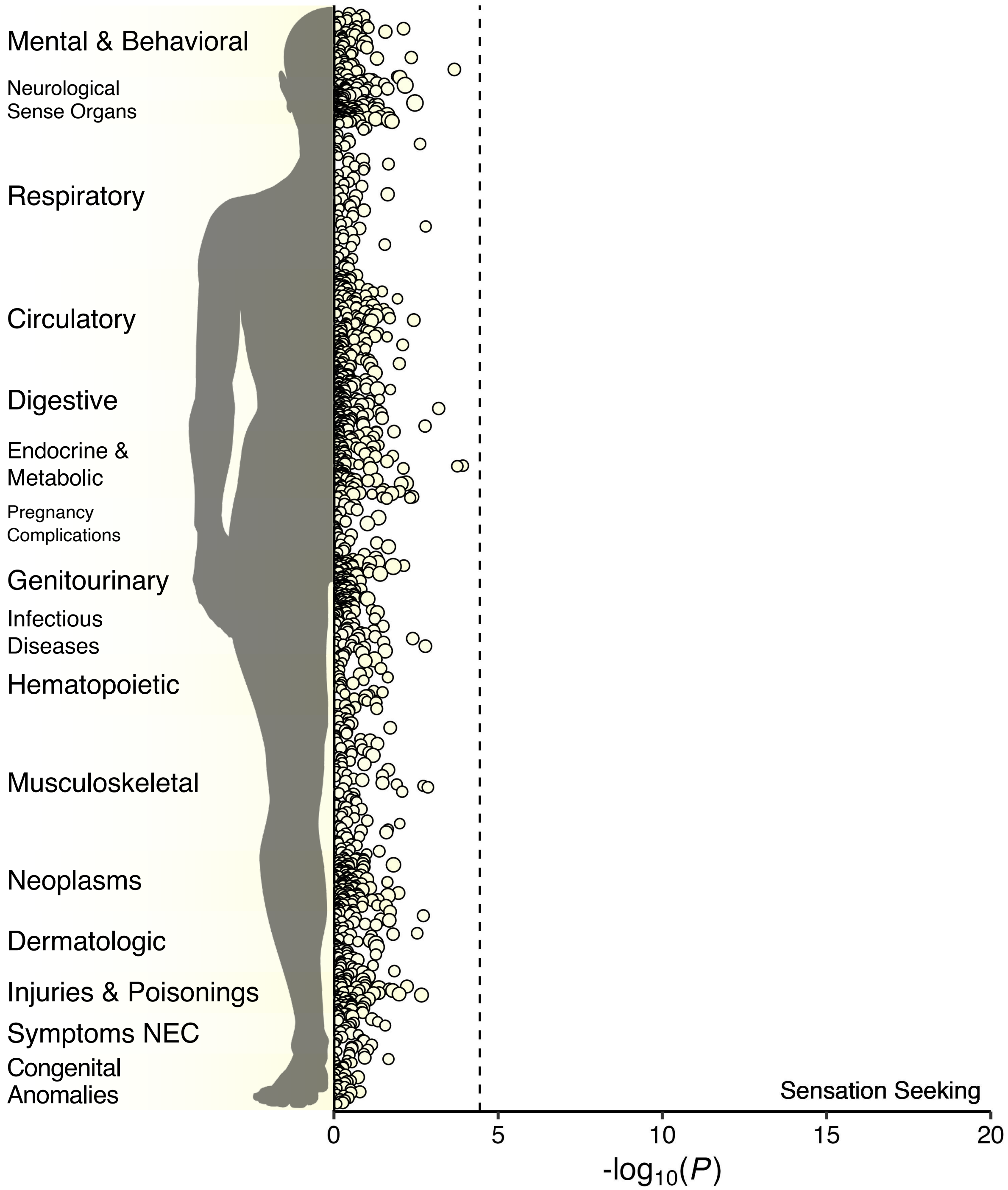


**Figure S20. Results of phenome-wide association study of sensation seeking polygenic score in BioVU.** Manhattan plot of the associations between the sensation seeking polygenic score and medical outcomes in BioVU. The y-axis refers to the category of medical outcome (or "phecode"), the x-axis refers to the statistical significance of the association on $-\log_{10}$ scale, and the dashed line denotes the Bonferroni-adjusted significance threshold. Circles denote associations that are nonsignificant after correction, upward-facing triangles are significant positive associations, and downward-facing triangles are significant negative associations. Size reflects the effect size, with larger points reflecting a larger absolute effect. For plotting purposes, some labels have been shortened or omitted. NEC = not elsewhere classified.
